# Understanding Pain in Polycystic Ovary Syndrome: Health Risks and Treatment Effectiveness

**DOI:** 10.1101/2024.10.15.24315513

**Authors:** Tess Cherlin, Stephanie Mohammed, Samantha Strydesky, Sasha Ottey, Katherine Sherif, Shefali S. Verma

## Abstract

Polycystic ovary syndrome (PCOS) is a prevalent endocrine disorder in women, often accompanied by various symptoms including significant pain, such as dysmenorrhea, abdominal, and pelvic pain, which remains underexplored. This retrospective study examines electronic health records (EHR) data to assess the prevalence of pain in women with PCOS. Conducted on January 2026, using data from 120 Health Care Organizations within the TriNetX Global Network, the study involved 103,675,738 women from diverse racial backgrounds. The analysis focused on the prevalence of pain among women with PCOS, both overall and in those prescribed PCOS-related medications. Relative risk ratios (RR) were calculated for future health outcomes and stratified by self-reported race. The study found that 20.67% of women with PCOS experienced pain, with the highest prevalence among Black or African American (32.70%) and White (30.78%) populations. Both the PCOS and PCOS and Pain cohorts exhibited increased RR for various health conditions, with significant differences noted across racial groups for infertility, ovarian cysts, obesity, and respiratory diseases. Additionally, women with PCOS who were treated with PCOS-related medications showed a decrease in pain diagnoses following treatment. In conclusion, this study highlights the critical need to address pain in the diagnosis and management of PCOS due to its significant impact on patient health outcomes.

**Impact Statement:** Insufficient data exist on the prevalence of pain in women with a PCOS diagnosis, and its associations with future health outcomes. Among, 597,638 women with PCOS in the TriNextX Global Network, 20.67% have dysmenorrhea, abdominal, and pelvic pain. Women with PCOS and Pain are at increased risk for developing ovarian cysts, infertility, T2D, and fatty liver disease and are at further risk when stratified by self-reported race groups.

## Introduction

According to the World Health Organization (WHO), polycystic ovary syndrome (PCOS) affects approximately 8-13% of women of reproductive age, with an alarming 70% of affected individuals remaining undiagnosed globally (Organization, 28 June 2023). The assessment of PCOS has been substantiated by multiple guidelines (Azziz et al., 2016; Teede et al., 2010) and has undergone refinement since its initial description by Stein and Leventhal in 1935 (Stein & Leventhal, 1935). Standard diagnostic criteria have evolved through international efforts, including conferences convened by the National Institutes of Health (NIH) in 1990 (Zawadri, 1992), the ESHRE/ASRM-sponsored PCOS consensus workshop group in Rotterdam in 2003 and 2004 (ESHRE & Group, 2004), and the International Evidence-based Guideline for the Assessment and Management of Polycystic Ovary Syndrome in 2018, most recently updated in 2023 (Mousa & Tay, 2023; Teede et al., 2018).

Recommendations for assessing PCOS encompass a multifaceted approach, including the evaluation of irregular menstrual cycles, ovulatory dysfunction, biochemical and clinical hyperandrogenism, ultrasound findings, serum Anti-Mullerian Hormone (AMH) levels, and various other factors such as ethnic disparities, cardiovascular disease risk, menopausal status, impaired glucose tolerance, and risk of type 2 diabetes mellitus (T2DM) (Mousa & Tay, 2023; Teede et al., 2018). Additionally, screening and managing psychological manifestations, implementing lifestyle interventions, and adhering to pharmacological treatment principles are integral aspects of PCOS management (Mousa & Tay, 2023; Teede et al., 2018). While the diagnostic criteria for PCOS primarily focus on reproductive and metabolic manifestations, the substantial burden of pain experienced among women with PCOS is a critical factor that warrants effective prevention and management of the disease.

The assessment of pain in PCOS necessitates a multidimensional approach, incorporating self-reported scales, clinical evaluation, and possibly imaging techniques to elucidate the underlying etiology and severity. Several commonly utilized assessment tools incorporate evaluations of pain among women with PCOS. The Polycystic Ovary Syndrome Health-Related Quality of Life Questionnaire (PCOSQ), developed by Cronin et al. in 1998, assesses various domains, including painful menstrual cycles (Cronin et al., 1998). Additionally, the SF-36 scale, examines eight dimensions of health, including bodily pain (McHorney et al., 1993). Women with PCOS across diverse demographic backgrounds have consistently reported lower SF-36 scores, specifically in the domain of bodily pain (Drosdzol et al., 2007; Elsenbruch et al., 2003; Hahn et al., 2005; Li et al., 2011). Furthermore, the Menorrhagia Outcomes Questionnaire, developed by Lamping et al. in 1998, evaluates both heavy menstrual bleeding (HMB) and the associated pain (Lamping et al., 1998). Despite the validation of these instruments, they may not comprehensively capture key symptoms expressed by patients with PCOS, especially those related to dysmenorrhea, abdominal, or pelvic pain. Insufficient data exist to highlight the prevalence of pain reported by women both before and after a PCOS diagnosis, as well as any associations between this pain and the condition itself and its long-term effects. To address this gap in research, we propose an investigation utilizing health records to shed light on this underexplored aspect of PCOS.

Electronic health records (EHRs) have become indispensable for managing vast amounts of clinical data, including patient demographics, medical history, medications, allergies, laboratory test results, vital signs, and imaging reports, as well as genetic information obtained from patient genomes when available. Given that EHRs contain comprehensive information about patient care, including the progression of signs and symptoms, severity, comorbidities, and treatments, they provide invaluable resources for conducting large-scale retrospective studies. EHR-based studies have been particularly valuable in assessing the prevalence of conditions that are often underdiagnosed or misdiagnosed in women (Kruse et al., 2018; Maletzky et al., 2022; Penrod et al., 2023). The temporal aspect of clinical events, such as the onset of symptoms, treatment administration, and follow-up visits, can also be mined from EHRs, providing crucial insights into disease trajectories and treatment efficacy (Zhao et al., 2017).

Pain, particularly in the context of PCOS, remains an underexplored area of research. By leveraging EHR data we can identify women with PCOS who have reported dysmenorrhea, abdominal, and pelvic pain. The objective of this research is to use EHR and look at longitudinal data retrospectively to determine the pain reported by women with PCOS and to compare this to women without PCOS. The primary hypothesis of this study is that women with PCOS experience a higher prevalence to pain (including dysmenorrhea, abdominal pain, and pelvic pain) compared to women without PCOS, and this prevalence varies by racial groups. The hypothesis aims to investigate the prevalence of pain in women with and without pain. Our approach will provide insights into the relationship between pain symptoms and PCOS contributing to a better understanding of the condition and potentially improving patient care.

## Methods

### Study Design

The data used in this study was collected on January 9^th^, 2026, from the TriNetX Global Network, which provided access to electronic medical records (diagnoses, procedures, medications, laboratory values, genomic information) from approximately 1196,361,552 million patients from 172 healthcare organizations in 19 different countries. TriNetX has a rigorous quality control pipeline which can be found in the platforms, documentation. Briefly, EHR data is received from HCOs in CSV format. TriNetX maps the data to a standard and controlled set of clinical terminology. Demographics data are mapped to HL7 administrative standards, diagnoses are represented by ICD codes, procedures are represented by ICD and CPT codes, and medications are mapped to RxNorm ingredients. The data is then transformed into a proprietary data format. Data cleaning is performed and records that don’t meet the TriNetX quality standards are excluded. Quality checks are done for formatting and records with missing required data are excluded. TriNetX does not impute or estimate clinical values to fill gaps in patients’ records and there is no guarantee of data completeness.

### Cohort Definitions

A retrospective cohort analysis was conducted for patients with PCOS and patients with PCOS and Pain. Patients who were identified as cases (met inclusion and exclusion criteria) were compared to their respective controls. Description of inclusion and exclusion criteria for each cohort can be found in **Table 1 – Figure supplement 1**.

For the PCOS cohort, PCOS was defined as either having a PCOS diagnosis (ICD-10-CM E28.2), or an irregular menstruation (ICD-10-CM N92.6) and hirsutism (ICD-10-CM L68.0) diagnosis, or an irregular menstruation (ICD-10-CM N92.6) and androgen excess (ICD-10-CM E28.1) diagnosis. PCOS controls were defined as having a physical examination (ICD-10-CM Z00.0) and none of the PCOS case criteria. PCOS participants also had to satisfy stringent exclusion criteria to avoid confounders. Exclusion criteria for PCOS consisted of Maternal care for benign tumor of corpus uteri (ICD-10-CM O34.1), Leiomyoma of uterus (ICD-10-CM D25), endometriosis (ICD-10-CM N80), Polyp of corpus uteri (ICD-10-CM N84.0), female pelvic inflammatory disease unspecified (ICD-10-CM N73.9), hypothyroidism (ICD-10-CM E03.8, E03.9), hyperprolactinemia (ICD-10-CM E22.1), and adrenal hyperplasia (ICD-10-CM E27.8, Q89.1). Description of inclusion and exclusion criteria for each cohort can be found in **Table 1 - Figure supplement 1**.

For the PCOS and Pain cohort, PCOS was defined the same as above. PCOS and Pain cases were defined as patients with a PCOS case as well as being diagnosed for either abdominal and pelvic pain (ICD-10-CM R10) or dysmenorrhea (ICD-10-CM N94.6) ± three months from their first PCOS diagnosis. PCOS and Pain controls were defined as patients with PCOS but no pain diagnoses. Description of inclusion and exclusion criteria for each cohort can be found in **Table 1 - Figure supplement 1**.

To compare cohorts (cases / controls), the first documented encounter or PCOS (case / controls) or PCOS and Pain (case / controls) was defined as an “index event” in TriNetX. Index events are the specific dates a patient satisfies all selected cohort criteria. Baseline characteristics are all assessed *before* the index event while all health outcomes are assessed *after* the index event. **Figure 1 - Figure supplement 2** shows a graphical representation of this relationship among index events and outcomes specified in this study.

### Propensity Score Matching

The TriNetX platform uses a cohort matching method called 1:1 propensity score matching (Austin, 2011). For each cohort analysis, cases were matched on the following criteria: age at the index event, self-reported race, overweight, obesity, and other hyperalimentation (ICD-10-CM E65-E69) status, type 2 diabetes mellitus (T2D) (ICD-10-CM E11) status, essential (primary) hypertension (ICD-10-CM I10) status, and hyperlipidemia, unspecified (ICD-10-CM E78.5) status. Baseline conditions were assessed up to one day before the index event. **Figure 1** shows the number of patients in each case and control cohort both at baseline and after propensity score matching.

**Figure 1.**
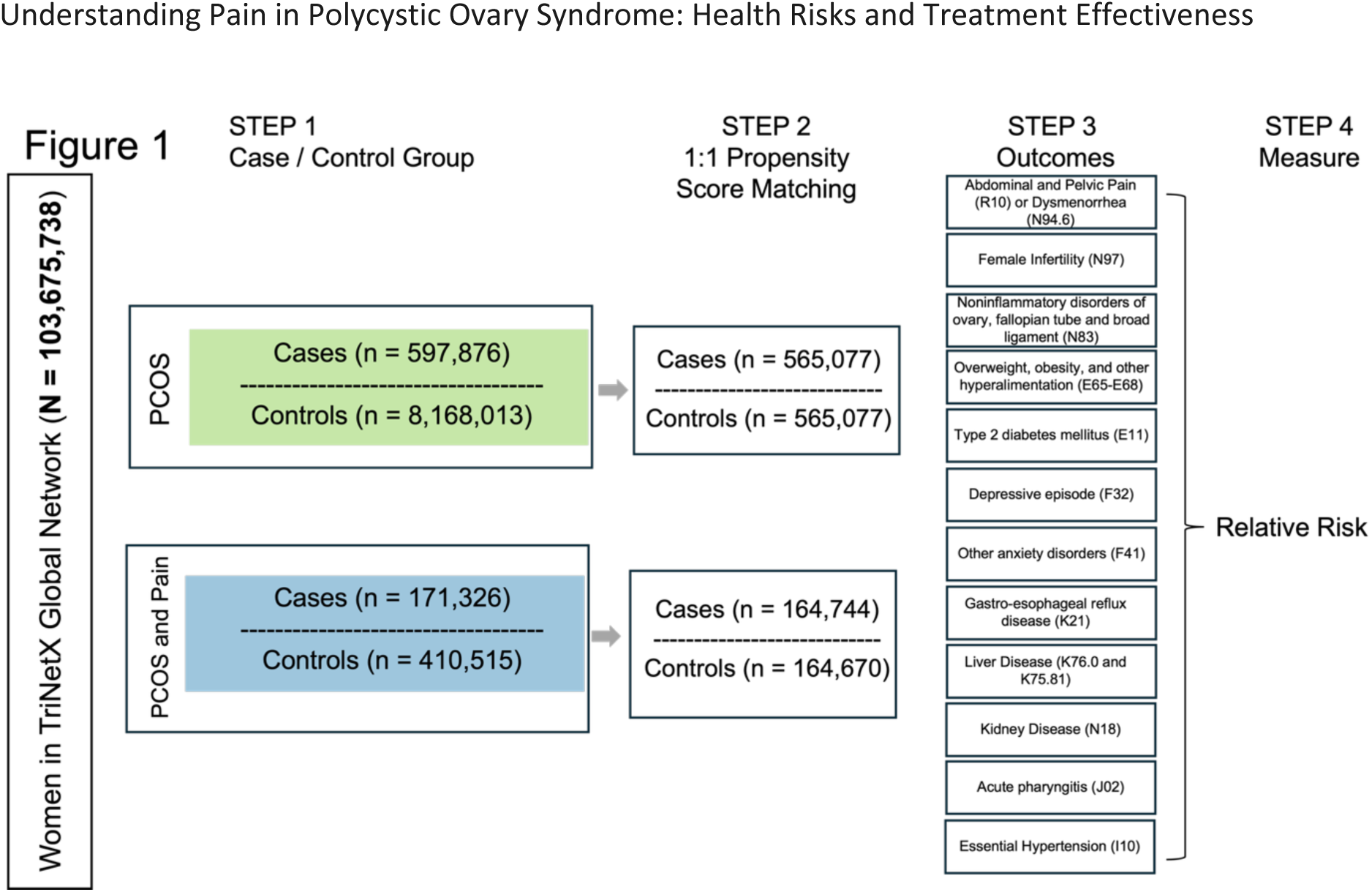
Analysis pipeline. Analysis pipeline to calculate relative risk ratios (RR) for future health outcomes in PCOS (green) and PCOS and Pain (blue) cohorts for the 103,675,738 women queried. STEP 1 shows the number of women in the case and controls for both the PCOS (green) and PCOS and Pain (blue) cohorts. STEP 2 shows the number of cases and controls after 1:1 propensity score matching. STEP 3 shows the different future health conditions that were considered for future health outcomes. STEP 4 shows that the final step is calculating the relative risk for the future health outcomes.

### Future Health Outcomes

For each of the cohorts listed above, we calculated relative risk ratios (RR) with 95% confidence intervals for 11 future health outcomes: abdominal and pelvic pain (R10) or dysmenorrhea (N94.6), female infertility (ICD-10-CM N97), noninflammatory disorders of ovary, fallopian tube and broad ligament (ICD-10-CM N83), obesity (ICD-10-CM E65-E68), Type 2 Diabetes (ICD-10-CM E11), depressive episode (ICD-10-CM F32), other anxiety disorders (F41), gastro-esophageal reflux disease (GERD) (ICD-10-CM K21), nonalcoholic steatohepatitis (ICD-10-CM K75.81) or fatty liver, not elsewhere classified (ICD-10-CM K76.0), chronic kidney disease (ICD-10-CM N18), and essential hypertension (ICD-10-CM I10). The RR for future health outcomes was calculated on participants who satisfied the 1:1 propensity score matching criteria (above). Differences in relative risks were calculated for significance by calculating the difference of two estimates (Altman & Bland, 2003). Future health outcomes were only considered if their first occurrence was at least 3 months after the index event. **Figure 1** shows the health outcomes assessed after 1:1 propensity score matching (Austin, 2011; Guo & Fraser, 2014; Haukoos & Lewis, 2015). These analyses were done within the TriNetX platform, and no individual-level data was extracted from the platform.

### Self-Reported Race Stratified Sub-Analysis

We did a follow-up analysis looking at health outcomes for patients with PCOS and PCOS and Pain compared to matched controls stratified by self-reported race and ethnicity. The following race categories are present in the TriNetX platform: American Indian or Alaskan Native, Asian, Black or African American, Native Hawaiian or Other Pacific Islander, Other, White, and Unknown Race. For our analysis, we included the following four race categories: Asian, Black or African American, and Other (American Indian or Alaskan Native or Native Hawaiian or Other Pacific Islander, or Other), and White **(Figure 2 - Figure supplement 3).** Due to the small population sizes of American Indian or Alaskan Native or Native Hawaiian or Other Pacific Islander, or Other, we decided to combine these four self-reported race groups together into one “Other” population. For the PCOS and Pain cohorts, we looked at ovarian cysts, infertility, obesity, T2D, depression, anxiety, GERD, pharyngitis, essential hypertension, liver disease, and kidney disease stratified by self-reported race (**Figure 2 - Figure supplement 3**).

### PCOS Medication Sub-Analysis

We performed a follow-up analysis looking at the number of patients with PCOS who were documented as having pain before being prescribed three common PCOS medications (systemic contraceptives (VA: HS200), metformin (RxNorm 6809), or spironolactone (RxNorm 9997)) as shown in **Figure 3 - Figure supplement 4**. Three cohorts were created on January 16^th^, 2026, in the TriNetX Global Network. There were approximately 190,888,407 million patients from 170 healthcare organizations in 19 different countries. Patients were included if they had a PCOS diagnosis (described above) *and* 1) ever had a systemic contraceptives prescription but not a metformin or spironolactone prescription, 2) ever had a metformin prescription but not a systemic contraceptives or spironolactone prescription, and 3) ever had a spironolactone but not a systemic contraceptives or metformin prescription. For patients with PCOS, we counted the number of participants who had a diagnosis code for either dysmenorrhea or abdominal and pelvic pain before the index event. The index event was defined as a participant having a PCOS diagnosis and a medication prescription at the same time. We then counted the number of patients with PCOS who reported either dysmenorrhea or abdominal and pelvic pain after the index event. We further compared the change in prevalence before and after the indexed event.

## Results

### Demographics of Women with PCOS in the TriNetX Global Network

We first identified participants with PCOS and associated comorbidities. The demographics and characteristics of both the PCOS and non-PCOS cohorts are detailed in **Table 2 - Figure supplement 5**. We queried the 103,675,738 women of any age from 172 Health Care Organizations (HCOs) in the TriNetX Global Network for PCOS and associated comorbidities. After applying stringent inclusion/exclusion criteria for PCOS and subsequent controls (see Methods) we found a 5.6% (n = 576,077) prevalence of PCOS at an average age of 28.1 (SD ± 9.04) in this population. When we stratified the PCOS participants in 10-year age groups, we observe that the majority of PCOS patients are either 21-30 years old (30.46%) or 31-40 years old (38.42%) **Figure 4 – Figure supplement 7.** Of those participants with PCOS, 4.85%, 13.15%, 23.32%, and 58.68% self-identified as Asian, Black or African American, Other, or White respectively (**Table 2 - Figure supplement 5**).

We then examined the prevalence of PCOS-associated comorbidities represented in the PCOS cohort. We observed that 16.28% of patient with PCOS had a diagnosis code for obesity, 5.93% had a diagnosis code for essential hypertension, 3.51% had a diagnosis code for T2D, and 2.96% had a diagnosis code for hyperlipidemia (**Table 2 - Figure supplement 5**). We also investigated the prevalence of other women’s health conditions related to PCOS such as infertility and ovarian cysts. **Table 2 - Figure supplement 5** shows that women with PCOS had a 2.34% prevalence of infertility and a 3.92% prevalence of ovarian cysts.

Since many women with PCOS are prescribed medications to help manage symptoms associated with the condition, we aimed to gain a deeper understanding of the prevalence of PCOS-prescribed medications in the PCOS cohort. We found that 14.88% of the PCOS cohort were prescribed systemic oral contraceptives, 6.82% were prescribed metformin, and 3.15% were prescribed spironolactone (**Table 2 - Figure supplement 5**).

A focus of our paper is understanding the impact pain has on women with PCOS; therefore, we looked at the prevalence of both dysmenorrhea and abdominal and pelvic pain. We observed that overall, there was a 20.67% prevalence of pain (2.85% prevalence of dysmenorrhea, 17.82% prevalence of abdominal or pelvic pain) (**Table 2 - Figure supplement 5**).

### Demographics of Women with PCOS and Pain in the TriNetX Global Network

As noted above, 20.67% of women with PCOS also had a pain diagnosis, encompassing either dysmenorrhea or pelvic and abdominal pain. We first examined the demographics of the study participants using data from the TriNetX Global Network. **Table 3 - Figure supplement 6** shows the comprehensive demographic results, highlighting the distribution of pain diagnoses for age, race, and other relevant demographic factors. This table provides a clear view of the demographic characteristics and their potential influence on the prevalence of pain among individuals with PCOS. Similar to the PCOS cohort, participants with PCOS and Pain had an average age of 28.5 (SD ± 8.78) with the majority of participants being between 21-30 years old (32.74%) or 31-40 years old (38.97%). Interestingly, the prevalence of PCOS and Pain was slightly higher at these ages compared to the prevalence of PCOS alone (**Figure 4 – Figure supplement 7**). Among the women with PCOS and Pain, 3.58% women were Asian, 14.54% Black or African America, 20.63% categorized as Other, and 61.25% self-reported as White (**Table 3 - Figure supplement 6**). However, when we looked at the prevalence of PCOS and Pain compared to controls (PCOS without pain) within a self-reported race group, we observed that the highest prevalence of PCOS and Pain was 48.11% in the Black or African American population followed by 44.76% in the Other population and 43.84% in the White population, and 27.20% in the Asian population. (**Table 4 - Figure supplement 8**). With respect to PCOS comorbidities, the cohort of individuals with both PCOS and Pain exhibited a higher prevalence of comorbid conditions compared to the entire population of individuals with PCOS. Specifically, 33.88% of PCOS and Pain participants had an obesity diagnosis, 12.43% had an essential hypertension diagnosis, 7.84% had a T2D diagnosis, and 7.39% had a hyperlipidemia diagnosis. These high prevalences represent a respective increase of 16.28%, 5.93%, 3.52%, and 2.96% compared to all participants with PCOS. Notably, as illustrated in **Figure 2**, there is at least a two-fold increase in the prevalence of each comorbid condition among those with both PCOS and Pain. This substantial increase highlights the heightened risk and burden of comorbidities within the PCOS and Pain cohort.

**Figure 2.**
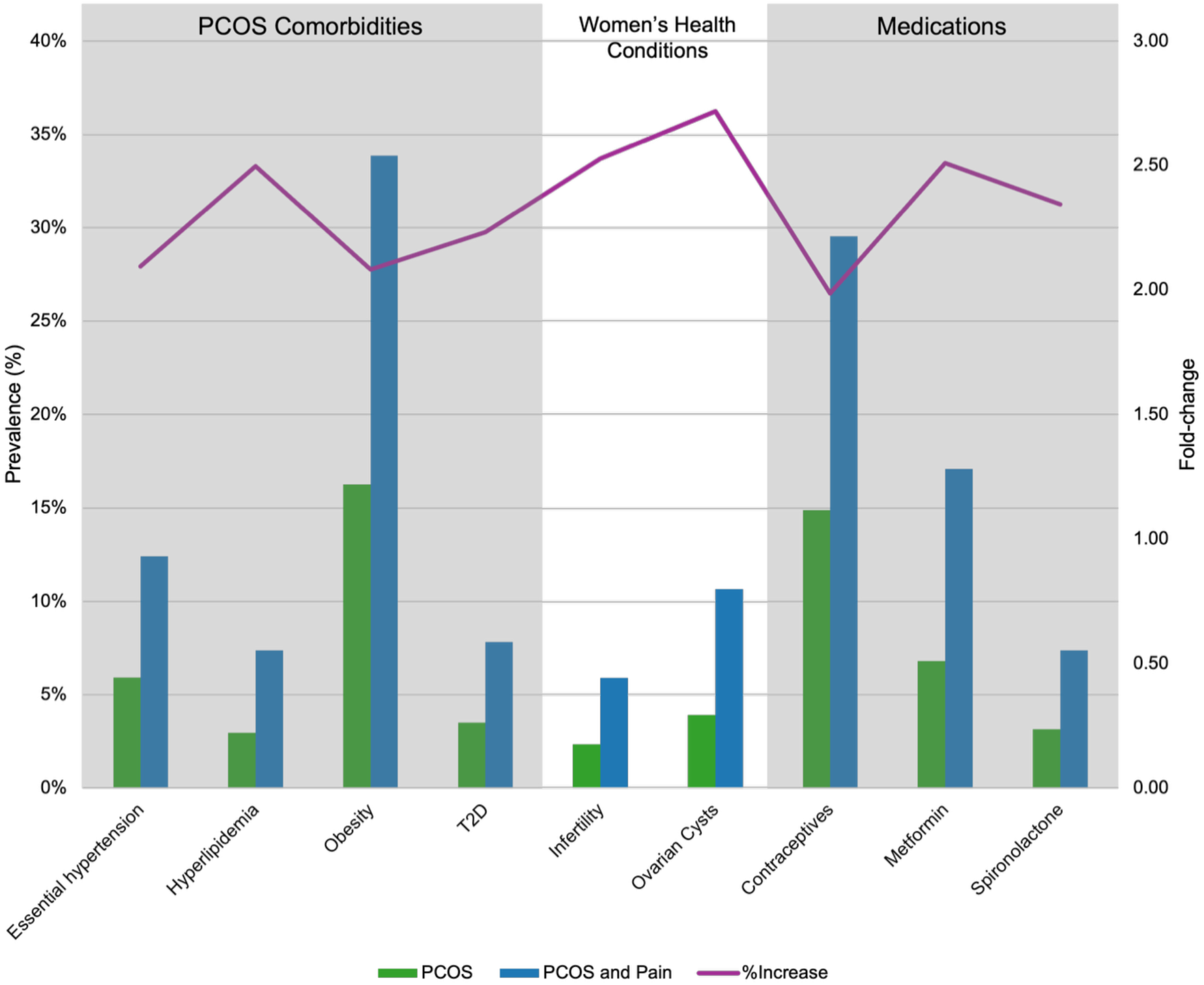
Prevalence of conditions and medications associated with PCOS and Pain. Barplots show the prevalence (%) (left y-axis) of different diseases associated with PCOS (green) and PCOS and Pain (blue) (x-axis). Purple line indicates the prevalence fold-change between the PCOS and PCOS and Pain cohorts (right y-axis).

We observed a similar trend with respect to two diseases affecting women with PCOS. We see a 2-fold increase in the prevalence of infertility (5.92%) and ovarian cysts (10.65%) in the PCOS and Pain cohort when we compared to the PCOS cohort as shown in **Figure 2**. Further, there is at least a 2-fold increase in prescriptions for all three common PCOS symptom-management medications in PCOS and Pain cohort compared to PCOS cohort **(Figure 2)**. **Risk of Future Health Outcomes for all PCOS vs. PCOS and Pain**

Given that PCOS symptoms first manifest in puberty and during reproductive years, we aimed to assess the risk for patients with PCOS, and those with both PCOS and Pain, developing future health outcomes. The risk (%) for the future health outcomes assessed in this study can be found in the risk column of **Table 5 – Figure supplement 9**. To start, we see that 21% of women with PCOS overall were at risk for a future diagnosis of Pain (abdominal and pelvic pain or dysmenorrhea). Of the comorbidities of PCOS, obesity, T2D, and essential hypertension had 20.7%, 5.1%, and 7.7% increased risk in the PCOS overall cohort and 20.4%, 5.2%, and 8.2% increased risk in the PCOS and Pain cohort respectively. Liver disease and kidney disease, which are on the rise in PCOS patients, were found to have a 4.0% and 0.7% increased risk in the PCOS cohort and at 5.4% and 0.9% increased risk in the PCOS and Pain cohort. The “Explore Outcome” feature on the TriNetX platform revealed that anxiety, depression, gastroesophageal reflux disease (GERD), and acute pharyngitis were the most common future health outcomes for women diagnosed with PCOS and Pain. When we looked at risk for these future health outcomes, we observed that 17.1%, 11.5%, 10.5%, 10.0% of patients with PCOS and 20.1%, 13.7%, 13.5%, 13.3% of patients with PCOS and Pain were at risk of developing anxiety, depression, acute pharyngitis, and GERD respectively. Overall, besides obesity, patients with PCOS and Pain showed a higher risk for the investigated future health outcomes than PCOS alone. We explore the relationship of future health outcome risk further in the results below.

### Relative Risk of Future Health Outcomes for all PCOS vs. PCOS and Pain

Given that PCOS symptoms first manifest in puberty and during reproductive years, we aimed to assess the relative risk for patients with PCOS, and those with both PCOS and Pain, developing future health outcomes. We calculated the RR for matched PCOS patient cohorts with their respective controls and PCOS and Pain patient cohorts with their respective controls (see Methods). Results are visualized in **Figure 3** and cohorts counts, RRs, and p-values for differences in risk ratios are provided in **Table 5 - Figure supplement 9**.

**Figure 3.**
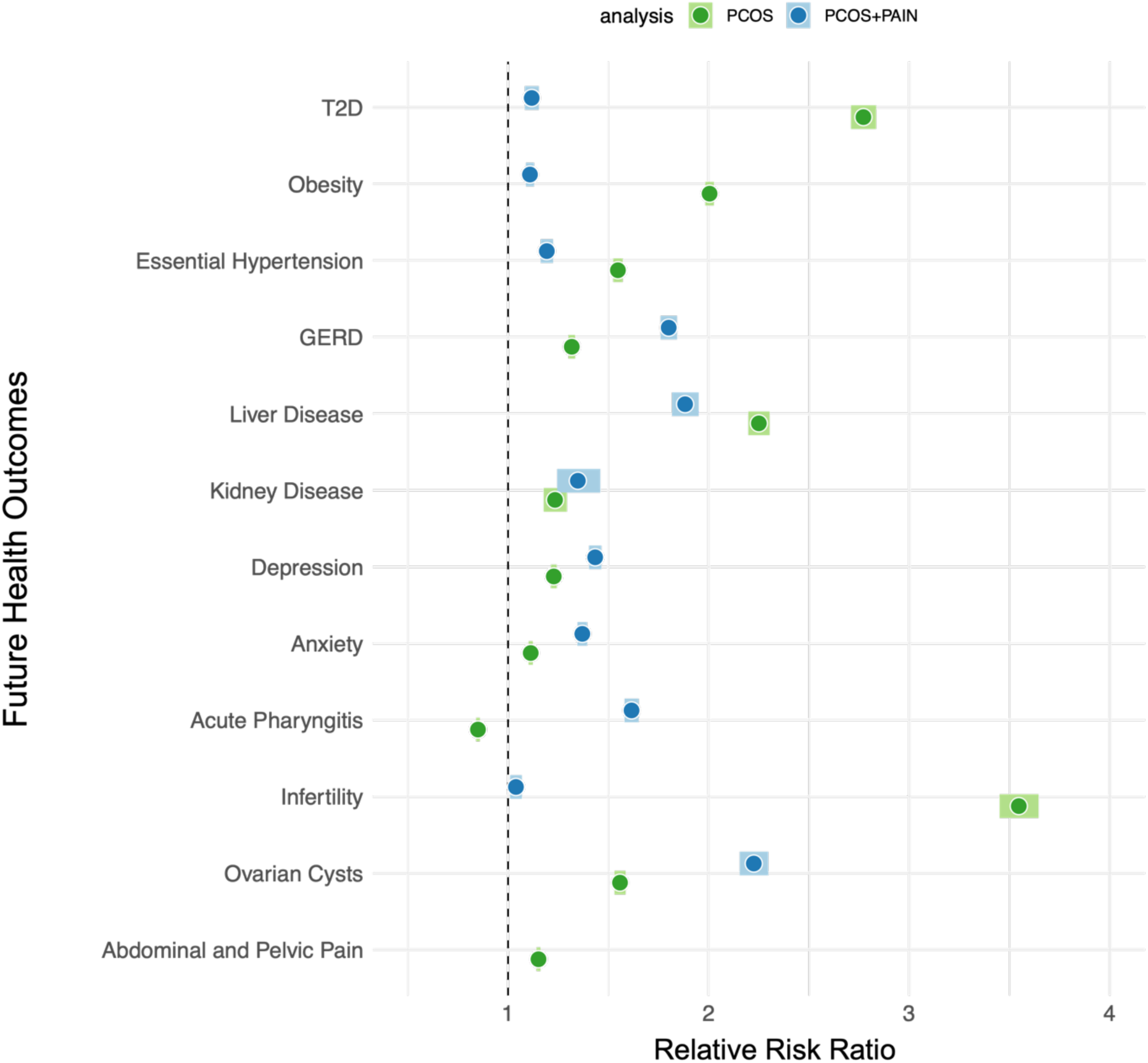
Relative risk ratios for future health outcomes associated with PCOS and Pain. Relative risk ratios (RR) (x-axis) for future health outcomes (y-axis) for both PCOS (green) and PCOS and Pain (blue) cohorts. Darker hued circles indicate RR, while lighter hued boxes indicate the 95% confidence intervals. The black dashed line is set 1 and is the threshold for RR, where > 1 is increased RR and < 1 is decreased RR.

In **Figure 3**, we see the RR for both participants within the PCOS cohort (green) for 12 health outcomes and PCOS and Pain cohort (blue) for 11 health outcomes. Aside from acute pharyngitis, the PCOS cohort has significantly increased risk for developing the following future health outcomes compared to matched controls: T2D, obesity, essential hypertension, GERD, liver disease, kidney disease, depression, anxiety, infertility, ovarian cysts, and pain. The PCOS and Pain cohort meanwhile has significantly increased risk for developing all of the following future health outcomes compared to their matched controls: T2D, obesity, essential hypertension, GERD, kidney disease, depression, anxiety, acute pharyngitis, infertility, ovarian cysts, and pain.

While almost all of the RR are increased for case cohorts compared to match controls, a few results stand out as being particularly interesting. Infertility, for example, a common complication associated with PCOS, has a RR of 3.55 (95% CI 3.45-3.64) in PCOS cases overall. Meanwhile, the RR for a future outcome of infertility for women with PCOS and Pain is a near-insignificant 1.04 (95% CI 1.04-1.07). This difference in relative risks has a p-value of 6.04E-820 (**Table 5 - Figure supplement 9**). On the other hand, ovarian cysts have a RR of 1.56 (95% CI 1.53-1.59) in PCOS cases overall, but an even higher RR in PCOS and Pain cases (RR=2.23, 95% CI 2.16-2.30). This relative risk difference is also statistically significant (p-value = 2.24E-36) (**Table 5 - Figure supplement 9**). In GERD, depression, anxiety, and acute pharyngitis all had higher RRs in PCOS and Pain cases vs. controls compared to PCOS cohort cases compared to controls (**Figure 3, Table 5 - Figure supplement 9**). We see that GERD, acute pharyngitis, depression, and anxiety are 1.80 (95% CI 1.76-1.84), 1.62 (95% CI 1.58-1.65), 1.43 (95% CI 1.40-1.47), and 1.37 (95% CI 1.35-1.39) in the PCOS and Pain cohort compared to 1.32 (95% CI 1.30-1.33), 0.85 (95% CI 0.84-0.86), 1.23 (95% CI 1.21-1.24) and 1.11 (95% CI 1.10-1.12) in the entire PCOS cohort at statistical significance **(Figure 3, Table 5 - Figure supplement 9**).

PCOS and PCOS and Pain cohorts were at comparable increased risk for developing future kidney disease. The RR of the PCOS cohort was 1.23 (95% CI 1.18-1.29) while the RR for the PCOS and Pain cohort was 1.35 (95% CI 1.24-1.46) **(Figure 3)**. The relative risk difference was not statistically significant (p-value = 6.64E-02) **(Table 5 - Figure supplement 9)**.

Obesity, T2D, essential hypertension, and liver disease had increased risk for the PCOS cohort compared to the PCOS and Pain cohort. T2D had a RR of 2.77 (95% CI 2.71-2.84) in the PCOS cohort compared to 1.12 (95% CI 1.08-1.15) in the PCOS in Pain cohort. Obesity had a RR of 2.00 (95% CI 1.98-2.03) in the PCOS cohort compared to 1.11 (95% CI 1.09-1.13) in the PCOS and Pain cohort. Meanwhile, the RR for liver disease was 2.25 (95% CI 2.20-2.30) in the PCOS cohort compared to 1.89 (95% CI 1.82-1.95) in the PCOS and Pain cohort. Finally, essential hypertension had a RR of 1.55 (95% CI 1.52-1.57) in the PCOS cohort and 1.19 (95% CI 1.16-1.22) in the PCOS and Pain cohort **(Figure 3)**. All relative risk differences were statistically significant **(Table 5 - Figure supplement 9)**.

### Relative Risk of Future Health Outcomes Stratified by Self-Reported Race

Since we were interested in the impact of Pain for women with PCOS, we next investigated if there were any race-specific risks (self-reported from EHR) for these outcomes. We stratified the PCOS and Pain case and control cohorts by self-reported race and calculated RR for the same 11 future health outcomes (methods) discussed above. **Figure 4(A-K)** shows the RR for each of the 11 future health outcomes in the Asian (orange), Black or African American (yellow), Other (red), and White (purple) PCOS and Pain cohorts. We observed significant race-specific differences in RR for a number of future health outcomes. **Figure 4A** shows that infertility has increased RR in the Other cohort (RR = 1.35, 95% CI 1.18-1.54) and Black or African American (RR = 1.23, 95% CI 1.13-1.34). Both the Other and Black or African American PCOS and Pain cohorts had significantly increased risk when compared to the Asian and White cohorts (adjusted p-value ≤ 0.005) (**Figure 4A, Table 6 - Figure supplement 10, Table 7 - Figure supplement 11)**. Ovarian cysts had increased RR across Asian (RR = 1.63, 95% CI 1.35-1.98), Black or African American (RR = 2.28, 95% CI 2.11-2.47), Other (RR = 2.31, 95% CI 2.06-2.60), and White (RR = 2.15, 95% CI 2.06-2.23) PCOS and Pain cohorts. However, there was a significantly increased RR for ovarian cysts in the Other, Black of African American, and White PCOS and Pain cohorts compared to the Asian PCOS and Pain cohort (adjusted p-value ≤ 0.05) (**Figure 4B**). Meanwhile, while all self-reported race cohorts show at least a 1.42 increased RR for depression, there was a significantly increased RR in Black or African American and Other PCOS and Pain cohorts compared to White PCOS and Pain cohorts (adjusted p-value ≤ 0.05) (**Figure 4F**). Interestingly, the Asian PCOS and Pain cohort had a decreased RR (RR = 0.8, 95% CI 0.51-1.27) for kidney disease compared to an increased RR in the Black or African American (RR = 1.53, 95% CI 1.45-1.61), Other (RR = 1.58, 95% CI 1.39-1.46), and White (RR = 1.42, 95% CI 1.39-1.46) PCOS and Pain cohorts (**Figure 4I)**. Moreover, here was a significant increased RR for kidney disease in the White PCOS and Pain cohort compared to the Asian PCOS and Pain cohort (adjusted p-value ≤ 0.05) (**Figure 4I)**. None of the PCOS and Pain self-reported race cohorts showed notable RR for the future health outcomes of T2D, Obesity, or Essential Hypertension (**Figure 4C-E**). On the other hand, all PCOS and Pain self-reported race groups had increased RR for anxiety (RR at least 1.36, 95% CI 1.33-1.39), GERD (RR at least 1.69, 95% CI 1.59-1.78), and acute pharyngitis (RR at least 1.48, 95% CI 1.30-1.69) (**Figure 4 G, J, K**).

**Figure 4.**
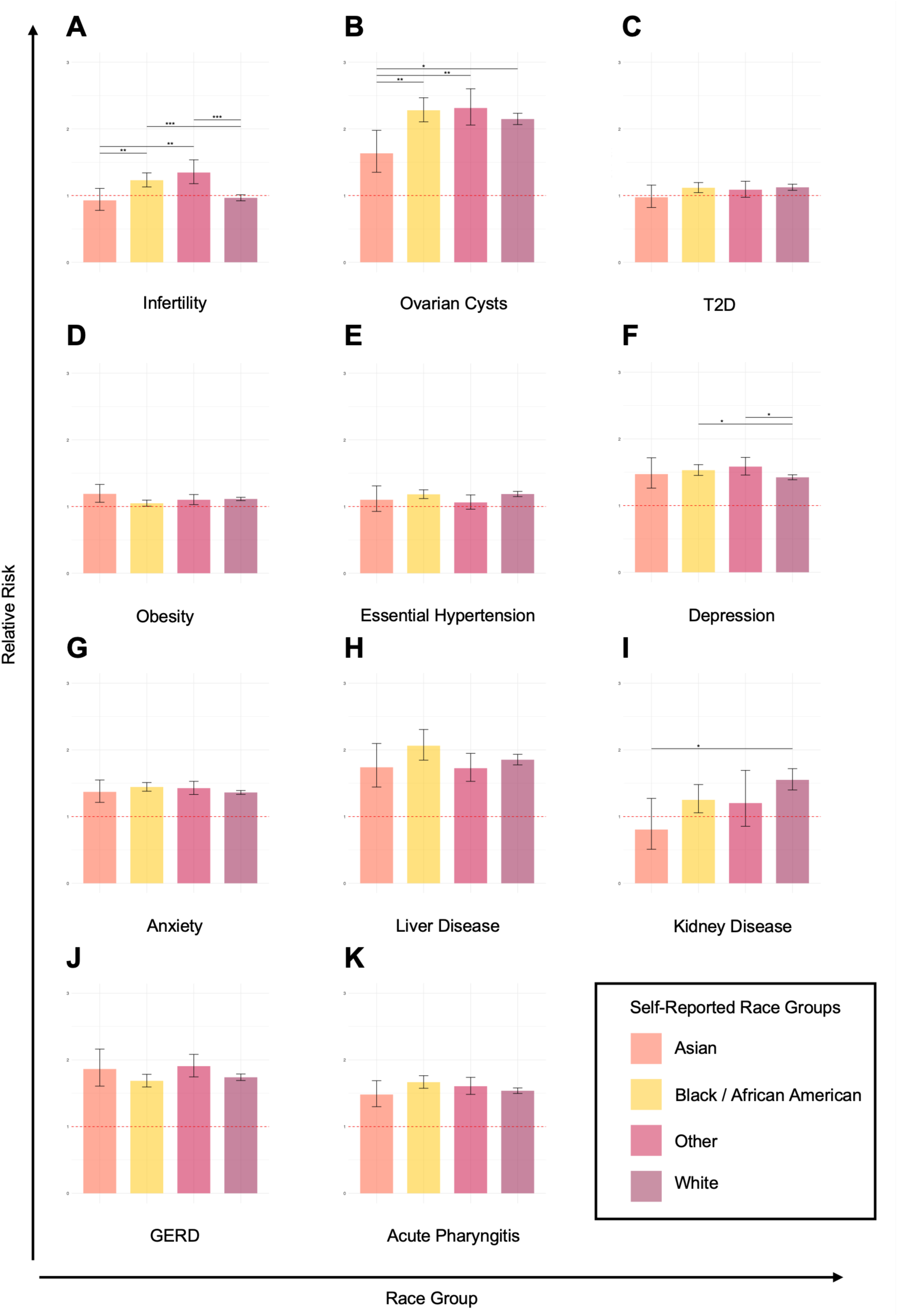
Parts A-K. Self-reported race-stratified relative risk ratios for future health outcomes. Relative risk ratios (RR) for future health outcomes (y-axis) stratified by self-reported race. Colors represent different self-reported race groups (x-axis): Asian (orange), Black or African American (yellow), Other (red), White (purple). Error bars represent the 95% confidence intervals. Significant differences between RR are represented by asterisks (*), where p-value ≤ 0.05 = *, p-value ≤ 0.005 = **, p-value ≤ 0.0005 = ***, and p-value ≤ 0.00005 = ****. Red dashed lines is set 1 and is the threshold for RR, where > 1 is increased RR and < 1 is decreased RR.

### Medications Prescribed to Patients with PCOS May Modify Pain Prevalence

Since women with PCOS are often prescribed medications to help their PCOS symptoms, we aimed to investigate if there were any changes in the pain diagnoses after being prescribed systemic contraceptives (COCPs), metformin, or spironolactone. For patients with PCOS who were prescribed each of the three medications exclusively, we calculated the percent who reported dysmenorrhea or abdominal and pelvic pain both before and after the prescription (see Methods). We found that there were 118,144 women with PCOS who were prescribed systemic contraceptives, 65,162 prescribed metformin, and 15,460 prescribed spironolactone. The prevalence of abdominal and pelvic pain diagnosis was 6-8x greater than that of a dysmenorrhea diagnosis for PCOS participants before they were prescribed PCOS-related medications (**Figure 5**).

**Figure 5.**
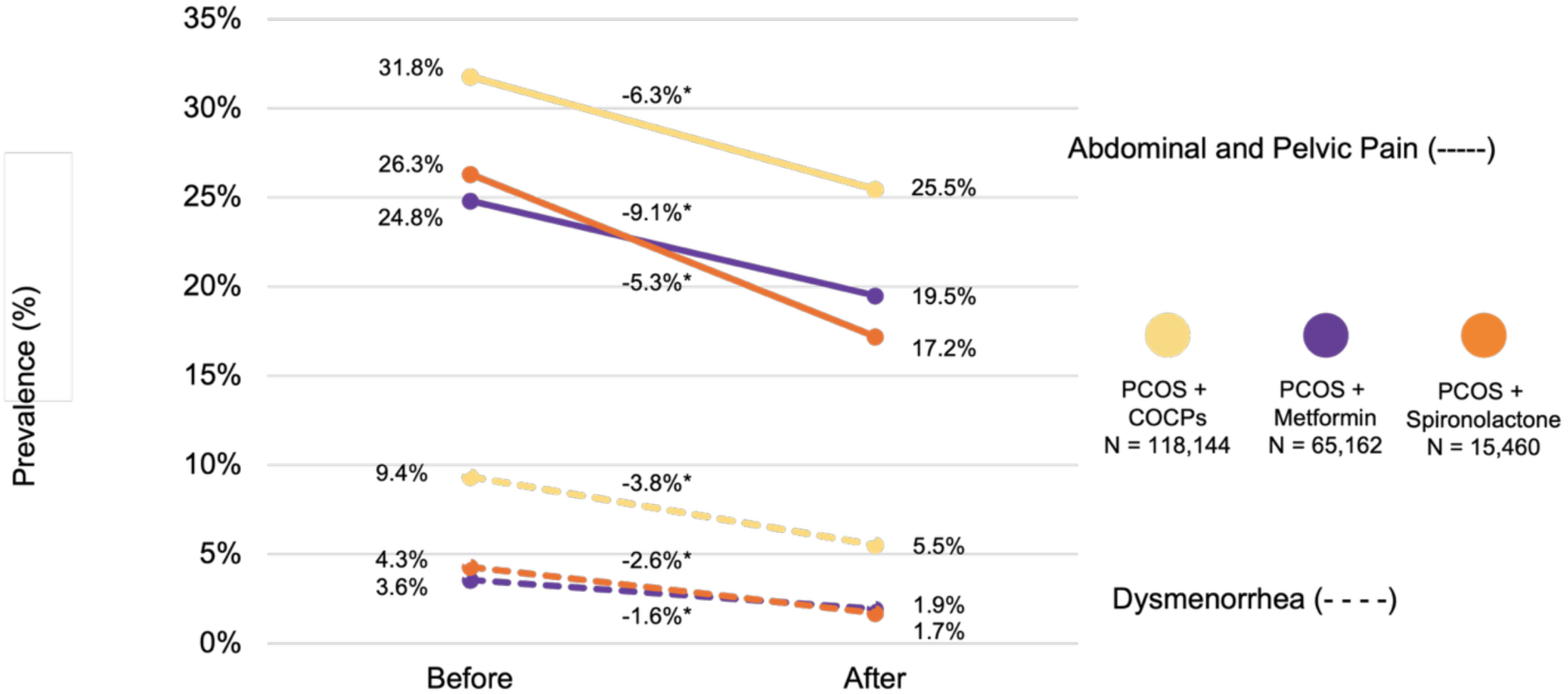
Prevalence of Pain for women with PCOS before and after medications. Prevalence (%) changes (y-axis) of pain for women with PCOS cohort before and after prescription of COCPs (yellow), metformin (purple), and spironolactone (orange) (x-axis). Analysis was done separately for abdominal and pelvic pain (solid lines) and dysmenorrhea (dashed lines).

Oftentimes, women with PCOS are prescribed COCPs, metformin, and spironolactone to manage their symptoms. We observed that women with PCOS had prescriptions for COCPs 17.8% of the time, metformin 10.3% of the time and spironolactone 2.3% of the time. As can be observed in **Figure 5**, participants with PCOS reported abdominal and pelvic pain at a prevalence of 31.8%, 24.8%, and 26.3% respectively before their first prescription of COCPs, metformin, and spironolactone. At least 3 months after being prescribed a PCOS-associated medication, we observe a significant reduction in the prevalence of abdominal and pelvic pain. Spironolactone shows the largest reduction of pain prevalence with a –9.1% reduction of pain diagnosis after prescription compared to before, followed by COCPs (−6.3%) and metformin (−5.3%). Similar results are observed for dysmenorrhea. While lower overall, there was also a decreased prevalence of pain for all three medications, the prevalence for dysmenorrhea was 9.4%, 3.6% and 4.3% for participants with PCOS prescribed COCPs, metformin, and spironolactone respectively. Unlike with abdominal and pelvic pain, COCP prescriptions were associated with the largest decrease in dysmenorrhea prevalence (−3.8%), followed by spironolactone (−2.6%), and (−1.6%).

## Discussion

Polycystic ovary syndrome is the most prevalent endocrine disorder among women (Mousa & Tay, 2023; Walters et al., 2018). Diagnosis and treatment plans are customized based on the symptoms presented by women. However, an important yet often overlooked variable is pain, which may manifest as dysmenorrhea, abdominal, or pelvic pain. The use of EHR data has facilitated access to patient records containing longitudinal clinical information, utilizing the readily available International Classification of Diseases (ICD) codes (Wu et al., 2016). Our study aimed to elucidate the prevalence and impact of pain among individuals with PCOS, as well as to investigate the relative risk of future health outcomes and the effectiveness of commonly prescribed medications on pain. Firstly, we observed a significantly higher prevalence of pain among women with PCOS compared to those without the condition. Specifically, 20.67% of women with PCOS reported experiencing pain, compared to 15.7% in the non-PCOS cohort. This increased prevalence reveals the substantial burden of pain as a symptom of PCOS, which often goes underreported and undertreated. Our demographic analysis of women with PCOS and Pain also revealed a difference in diagnosis of pain across self-reported race groups and was especially high in the Black or African American population (32.7%) and White population (30.78%). These findings suggest that pain is a significant symptom of PCOS that can vary across different demographic groups. The high prevalence of pain underscores the need for healthcare clinicians to routinely assess and address pain in the management of PCOS, particularly in racially diverse populations. The diversity in pain perception and reporting among different racial groups can be influenced by a variety of factors, including genetic differences, cultural attitude towards pain, access to healthcare, and socioeconomic status. Women of different racial groups often experience different severities in pain (Portenoy et al., 2004). This can lead to disparities in pain management and treatment outcomes (Campbell & Edwards, 2012; Jamieson & Steege, 1996). Additionally, cultural differences may also affect how individuals report pain and their willingness to seek medical help (Hadjiconstantinou et al., 2017).

PCOS manifests with many other concomitant conditions (Anagnostis et al., 2018; Asuncion et al., 2000; Balen et al., 2016; Escobar-Morreale et al., 2011; Hadjiconstantinou et al., 2017; Kitzinger & Willmott, 2002; Patel, 2018). Our study revealed that women with PCOS and Pain have at least a 2-fold increased prevalence of other health conditions at baseline compared to women with PCOS in general. The prevalence of obesity in the PCOS and Pain cohort was 33.88% compared to a 14.63% prevalence in the entire PCOS cohort. Excess abdominal visceral fat is well-documented to increase inflammation (Després, 2012) and PCOS is considered a pro-inflammatory condition linked with cardiovascular disease (CVD) and T2D. This inflammation, in turn, can underlie obesity, CVD, and insulin resistance (IR) (Abraham Gnanadass et al., 2021; Osborn & Olefsky, 2012). Our data show that 12.43% of patients with PCOS and Pain also had a diagnosis for essential hypertension and 7.84% of PCOS and Pain patients had a T2D diagnosis. These results underscore the health challenges faced by individuals dealing with both PCOS and Pain issues necessitating treatment approaches that address both the syndrome itself and its accompanying symptoms.

Women with PCOS are significantly at risk for future health outcomes such as infertility, T2D, coronary heart disease, dyslipidemia, depression, non-alcoholic fatty liver disease, and obstructive sleep (Anagnostis et al., 2018; Ávila et al., 2014; Chaudhuri, 2023; McGowan, 2011; Patel, 2018; Zore et al., 2017). Our results also highlight specific risks for different subgroups (PCOS overall and PCOS and Pain). In the overall PCOS cohort, the highest future health outcome risks are for infertility (RR = 3.54) and T2D (RR = 2.77). However, in patients with PCOS and Pain, the highest risks are for ovarian cysts (RR = 2.23). Ovarian cysts are a hallmark feature of polycystic ovarian morphology (PCOM), which is caused by immature/arrested follicles that do not ovulate and cause a “string of pearls” appearance and enlarging of the ovaries (Adashi et al., 2023; Tsilchorozidou et al., 2004). Ovarian cysts have long been disputed by the PCOS research community as not being associated with PCOS and therefore, not being associated with pain. However, the magnitude of this risk as shown in our results underscores the importance of regular monitoring and appropriate management strategies for patients presenting with both PCOS and pain symptoms. Liver disease had high and comparable RR in both cohorts with overall PCOS (RR = 2.23) and PCOS and Pain (RR = 1.89). PCOS is known to be linked with non-alcoholic fatty liver disease (NAFLD) (Butt & Devi, 2024; Kumarendran et al., 2018; Torres & Harrison, 2016). This association between PCOS and pain and liver disease may be explained by the shared metabolic disturbances common to both PCOS and NAFLD, such as insulin resistance and dyslipidemia (Georgescu, 2022; Qu et al., 2013; Torres & Harrison, 2016). The presence of chronic pain could potentially exacerbate these metabolic imbalances through various mechanisms, including altered stress responses and lifestyle factors (Kivimäki et al., 2023). These findings suggest that patients with PCOS who also experience chronic pain may represent a distinct phenotype with unique risk profiles. In contrast, women with PCOS without documented pain demonstrated higher relative risks for infertility, obesity, and T2D, suggesting a more metabolically driven PCOS phenotype. The differing RR patterns between PCOS with and without pain may therefore reflect heterogeneity in underlying pathophysiology, symptom recognition, or healthcare utilization. Further longitudinal and mechanistic studies will be needed to better understand these distinct clinical trajectories. Additionally, the increased risk for future health conditions in the PCOS and Pain cohort also suggest that pain may be an important marker for identifying individuals at risk of developing future health outcomes, necessitating more vigilant monitoring and proactive intervention.

Women with PCOS had a higher future risk of depression (RR=1.23) and anxiety (RR=1.11). These associations were substantially stronger in the PCOS and Pain cohort (depression RR=1.43; anxiety RR=1.37). This pattern aligns with prior evidence and supports routine mental health screening as part of PCOS care (Teede et al., 2018; Teede et al., 2023). Clinically, the higher RR estimates in the pain-enriched PCOS subgroup can be supported by the understanding that persistent pain associated with PCOS (dysmenorrhea, abdominal and pelvic pain), which can amplify stress, sleep disruption, and functional impairment, all of which can worsen mood and anxiety and increase healthcare contacts where these diagnoses are captured (O’Brien & Bosak, 2025; Sai & Mahaparale, 2024). We also observed increased future GERD risk in the PCOS overall cohort (RR=1.32), with a marked elevation in the PCOS and Pain cohort (RR=1.80). The increased RR for GERD in women with PCOS and PCOS and Pain supports the established model that obesity and central adiposity in PCOS, particularly in those with pain, is a factor for GERD and its complications (Hampel et al., 2005). Finally, acute pharyngitis showed increased risk specifically in the PCOS and Pain cohort (RR=1.62). This may reflect reflux-related upper airway irritation (laryngopharyngeal reflux), which has been linked to chronic pharyngitis-type presentations, as well as utilization/coding effects in a subgroup of PCOS patients with more frequent clinical encounters (Cui et al., 2024).

Lastly, our analysis explored the impact of common PCOS medications on pain management. We found that prescriptions for COCPs, metformin, and spironolactone are associated with a reduction in reported pain symptoms. Specifically, individuals who received these medications showed a 5.00% average decreased prevalence of pain diagnoses after treatment, suggesting that these medications may also manage pain symptoms in individuals with PCOS. A recent publication looked at the association of PCOS-related medications with adverse drug reactions (ADRs) for women with PCOS and found that metformin and COCPs was significantly associated with abdominal pain (Sidra et al., 2019). However, this study did not measure the association of ADRs with pain before and after the prescription of PCOS medication. Our results offer insights for application showing that efficient pharmacological management of PCOS symptoms can also help alleviate associated pain. Additionally, the efficacy of these medications in reducing pain, specifically spironolactone and COCPs, which are prescribed in PCOS for their antiandrogenic effects, may suggest hyperandrogenism to be a contributor to increased pain in PCOS, and a potential target for addressing pain in PCOS. Furthermore, the advantages of these treatments may be beneficial, not just in managing typical PCOS symptoms, but also in tackling the significant burden of pain experienced by many women with PCOS, highlighting a valuable role in drug repurposing.

Our study has limitations that need to be considered when interpreting the results. Firstly, relying on ICD codes to identify pain and other health outcomes may not capture the range of experiences and clinical intricacies. While these codes offer an approach to data collection, they might not fully reflect variations in pain severity or the personal experiences of those, with PCOS (Kataria & Ravindran, 2020). Although extensive, the use of EHR data may still contain gaps or discrepancies that could impact the accuracy of our results (Madden et al., 2016). Furthermore, since this study is observational, by nature it cannot establish a causal relationship between PCOS, pain, and future health outcomes. Moreover, the demographic variations observed—especially the higher occurrence of pain among individuals—could be influenced by socio-economic factors, access to healthcare, nutrition, and other unmeasured variables. In addition, self-reported race was not captured the same globally as it is not a variable that is coded by HCOs world-wide. Lastly, TriNetX captures only medication prescriptions, which does not allow our analysis to consider adherence issues, dosage differences, or concurrent treatments that may influence the outcomes observed.

We were unable to evaluate the use of analgesics or anti-inflammatory medications, as over-the-counter pain medications commonly used for dysmenorrhea and pelvic pain are not consistently captured within the TriNextX electronic health record system. This limitation prevents the assessment of how pain-specific treatments may influence reported pain outcomes.

Future research should focus on overcoming these limitations through studies with detailed clinical assessments and a broader range of demographic and socio-economic factors.

## Conclusion

Various pain subtypes can profoundly affect the daily lives of PCOS patients. Due to limited research in clinical and laboratory settings, the effects and underlying mechanisms of pain remain unclear. Our study highlights the significant prevalence and impact of pain in women with PCOS, revealing critical differences across racial groups and underscoring the heightened risk for future health complications in those experiencing pain. These findings emphasize the importance of comprehensive pain assessment, management and inclusion as guidelines in the standard care of PCOS, with a particular focus on addressing racial disparities. Additionally, the observed effectiveness of medications such as systemic oral contraceptives, metformin, and spironolactone in reducing pain symptoms provide valuable insights for clinical practice, suggesting that these treatments can offer dual benefits in managing both PCOS and associated pain. Dysmenorrhea, abdominal, and pelvic pain are common experiences in women with PCOS, in the absence of pelvic-related conditions that can contribute to this type of pain, such as pelvic inflammatory disease, endometriosis, and fibroids. It is crucial to distinguish between pain originating from PCOS and Pain arising from comorbidities to ensure appropriate management and targeted treatment strategies for improving the quality of life in affected individuals.

## Authors’ Contributions

SSV, SO, and KS conceived and supervised the study. TC and SSV designed the methods. TC and SS performed the analysis. TC analyzed the data and designed the figures. SM and TC wrote the manuscript. All authors interpreted the results. All authors read, edited, and approved the final manuscript.

## Compliance with Ethical Standards

Not Applicable

## Data Sharing

All summary-level data has all been included in this manuscript.

## Data Availability

All summary-level data has all been included in this manuscript

## Acknowledgements

We would like to acknowledge the patients in the TriNetX Global Network without whom this work would not be possible.

## Supplemental Materials

**Table 1 – Figure Supplement 1.**
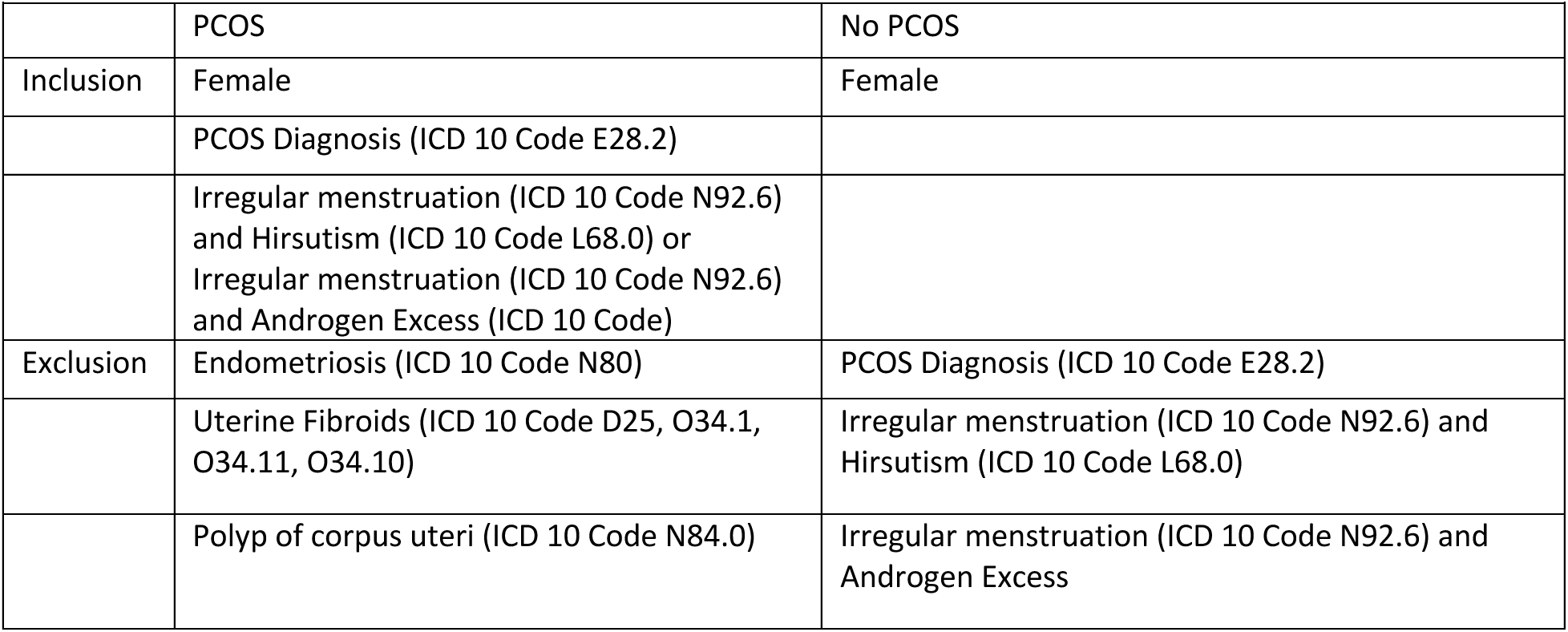
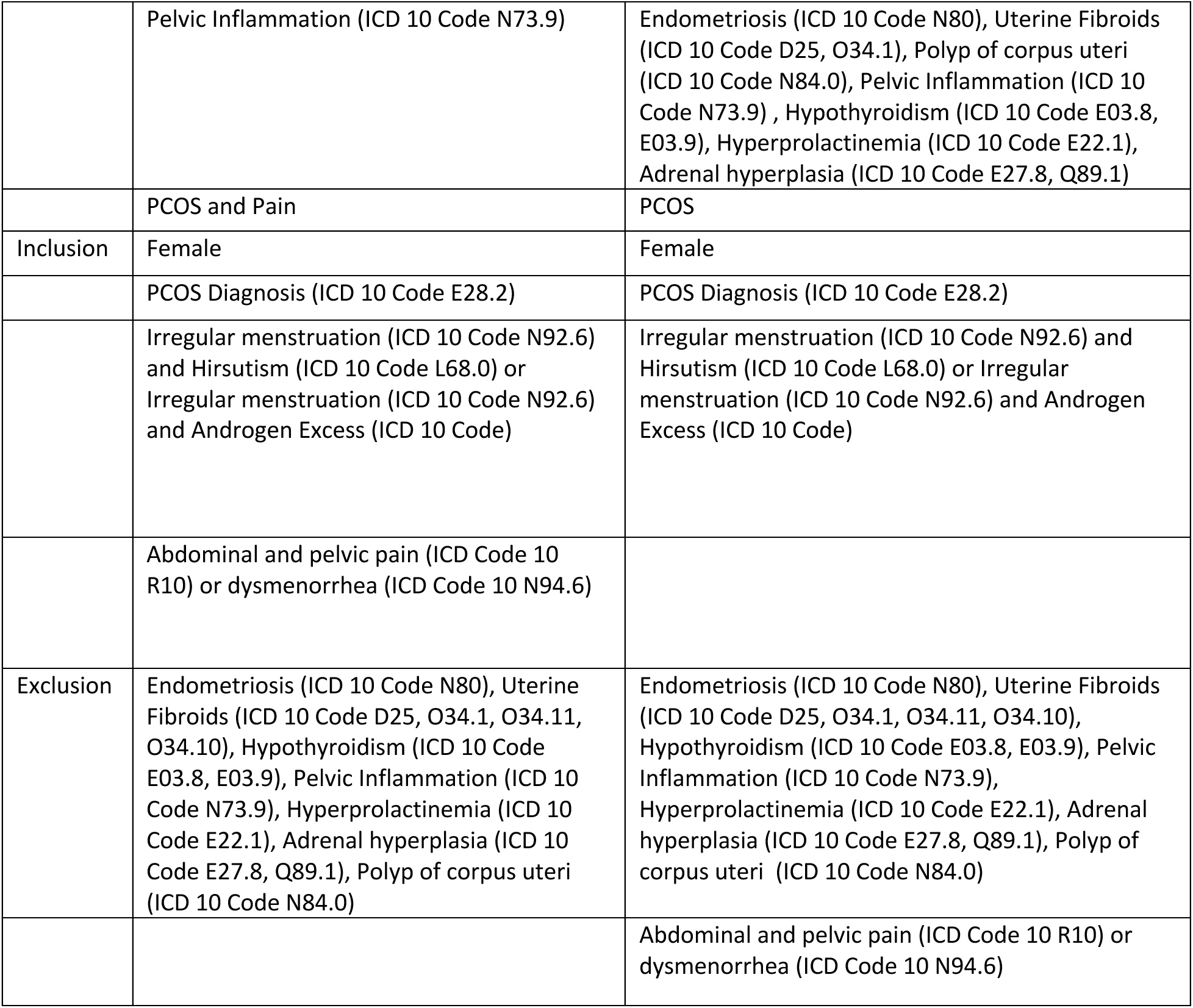
Inclusion and exclusion criteria for PCOS (top) and PCOS and Pain (bottom) cases and control cohorts.

|  |  |  |
| --- | --- | --- |
|  | PCOS | No PCOS |
| Inclusion | Female | Female |
|  | PCOS Diagnosis (ICD 10 Code E28.2) |  |
|  | Irregular menstruation (ICD 10 Code N92.6) and Hirsutism (ICD 10 Code L68.0) or Irregular menstruation (ICD 10 Code N92.6) and Androgen Excess (ICD 10 Code) |  |
| Exclusion | Endometriosis (ICD 10 Code N80) | PCOS Diagnosis (ICD 10 Code E28.2) |
|  | Uterine Fibroids (ICD 10 Code D25, O34.1, O34.11, O34.10) | Irregular menstruation (ICD 10 Code N92.6) and Hirsutism (ICD 10 Code L68.0) |
|  | Polyp of corpus uteri (ICD 10 Code N84.0) | Irregular menstruation (ICD 10 Code N92.6) and Androgen Excess |
|  | Pelvic Inflammation (ICD 10 Code N73.9) | Endometriosis (ICD 10 Code N80), Uterine Fibroids (ICD 10 Code D25, O34.1), Polyp of corpus uteri (ICD 10 Code N84.0), Pelvic Inflammation (ICD 10 Code N73.9) , Hypothyroidism (ICD 10 Code E03.8, E03.9), Hyperprolactinemia (ICD 10 Code E22.1), Adrenal hyperplasia (ICD 10 Code E27.8, Q89.1) |
|  | PCOS and Pain | PCOS |
| Inclusion | Female | Female |
|  | PCOS Diagnosis (ICD 10 Code E28.2) | PCOS Diagnosis (ICD 10 Code E28.2) |
|  | Irregular menstruation (ICD 10 Code N92.6) and Hirsutism (ICD 10 Code L68.0) or Irregular menstruation (ICD 10 Code N92.6) and Androgen Excess (ICD 10 Code) | Irregular menstruation (ICD 10 Code N92.6) and Hirsutism (ICD 10 Code L68.0) or Irregular menstruation (ICD 10 Code N92.6) and Androgen Excess (ICD 10 Code) |
|  | Abdominal and pelvic pain (ICD Code 10 R10) or dysmenorrhea (ICD Code 10 N94.6) |  |
| Exclusion | Endometriosis (ICD 10 Code N80), Uterine Fibroids (ICD 10 Code D25, O34.1, O34.11, O34.10), Hypothyroidism (ICD 10 Code E03.8, E03.9), Pelvic Inflammation (ICD 10 Code N73.9), Hyperprolactinemia (ICD 10 Code E22.1), Adrenal hyperplasia (ICD 10 Code E27.8, Q89.1), Polyp of corpus uteri (ICD 10 Code N84.0) | Endometriosis (ICD 10 Code N80), Uterine Fibroids (ICD 10 Code D25, O34.1, O34.11, O34.10), Hypothyroidism (ICD 10 Code E03.8, E03.9), Pelvic Inflammation (ICD 10 Code N73.9), Hyperprolactinemia (ICD 10 Code E22.1), Adrenal hyperplasia (ICD 10 Code E27.8, Q89.1), Polyp of corpus uteri (ICD 10 Code N84.0) |
|  |  | Abdominal and pelvic pain (ICD Code 10 R10) or dysmenorrhea (ICD Code 10 N94.6) |

**Figure 1 – Figure Supplement 2.**
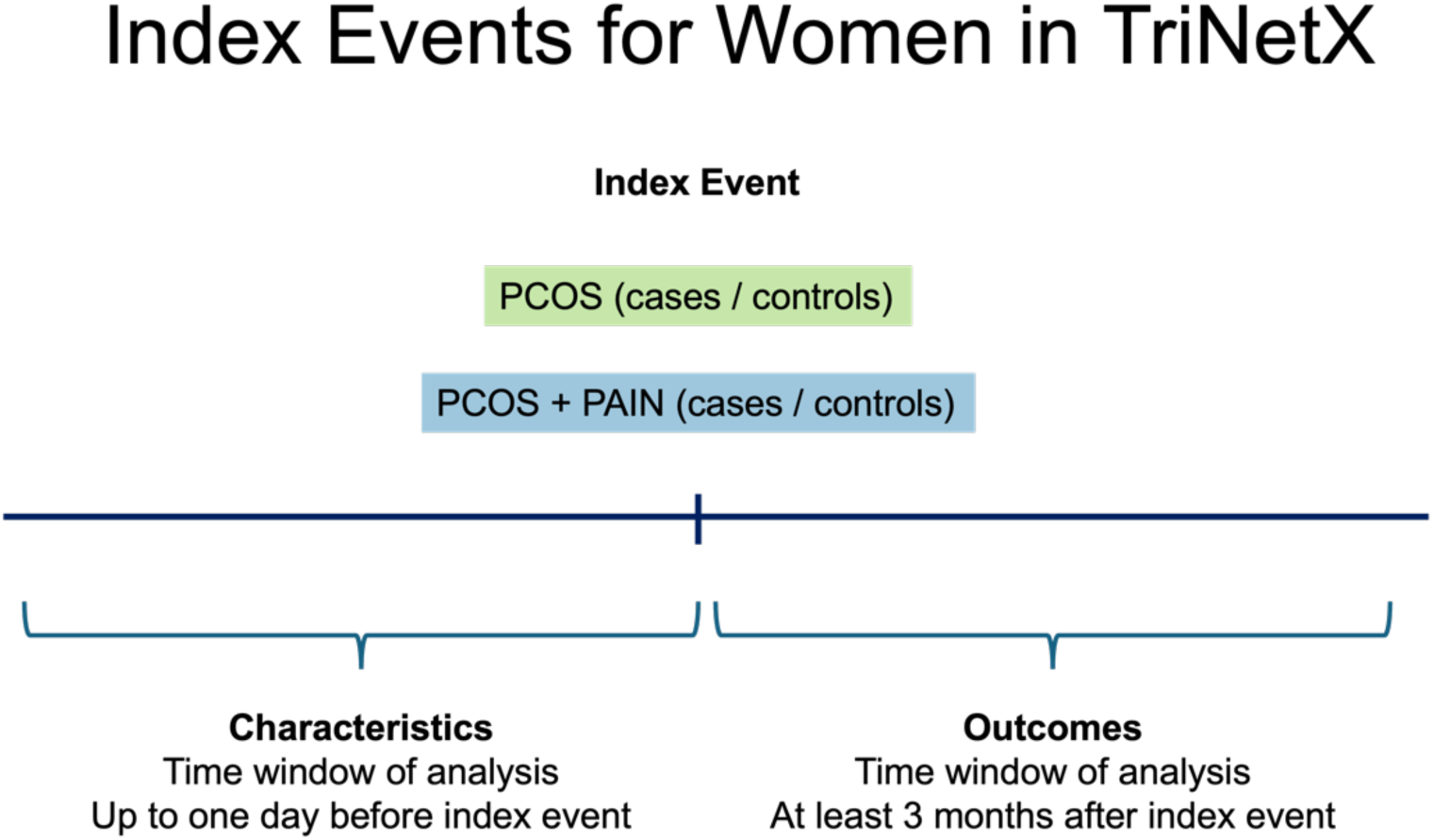
Index Events for Women in TriNetX. Schematic representing how events were indexed in TriNetX for both the PCOS (green) and PCOS and Pain (blue) cohorts.

**Figure 2 – Figure Supplement 3.**
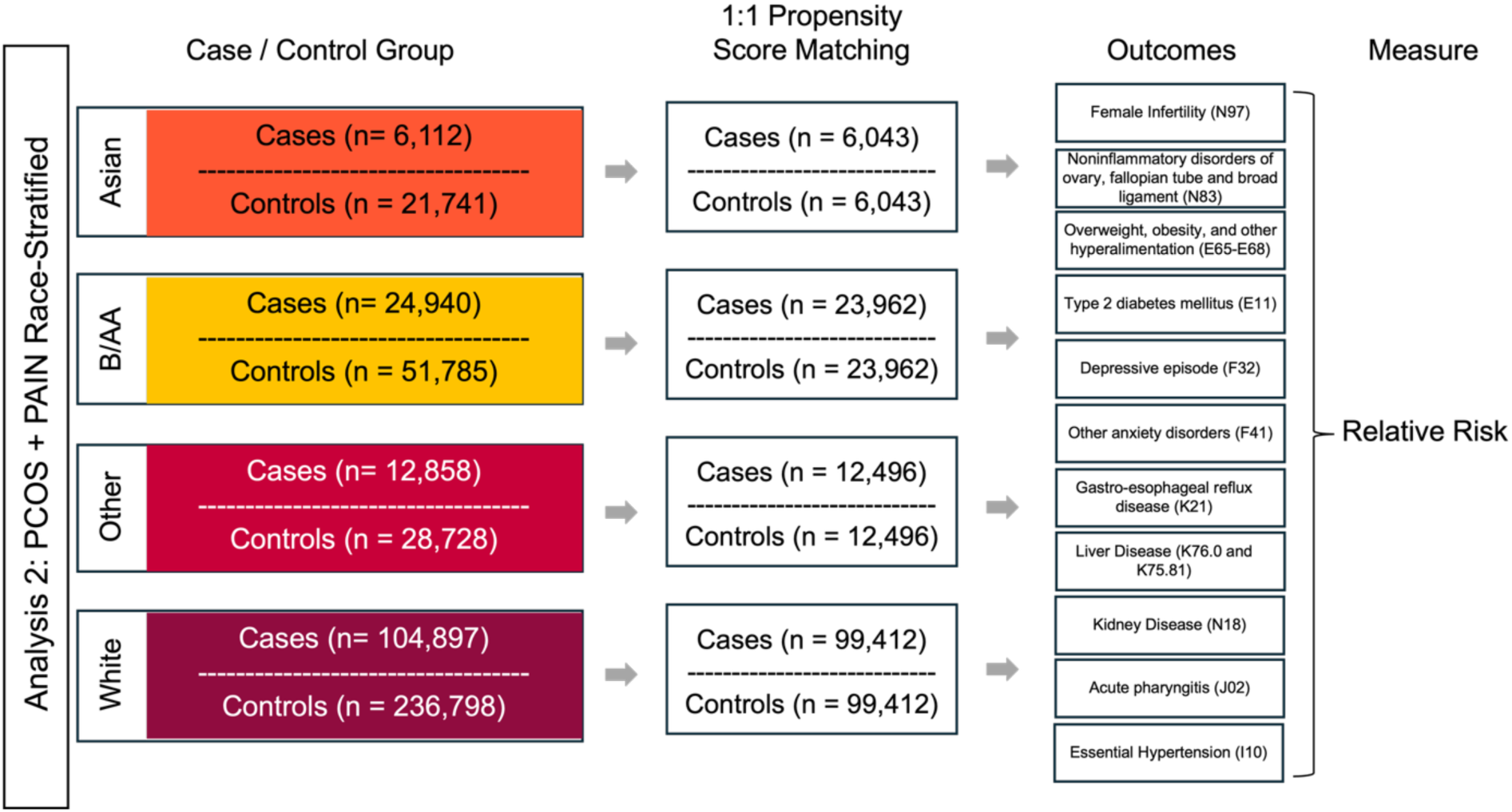
Analysis pipeline for self-reported race groups. Self-reported race-stratified TriNetX relative risk ratio analysis pipeline for future health outcomes in PCOS and Pain cohorts. Colors represent different self-reported race groups: Asian (orange), Black or African American (yellow), Other (red), White (purple).

**Figure 3 – Figure Supplement 4.**
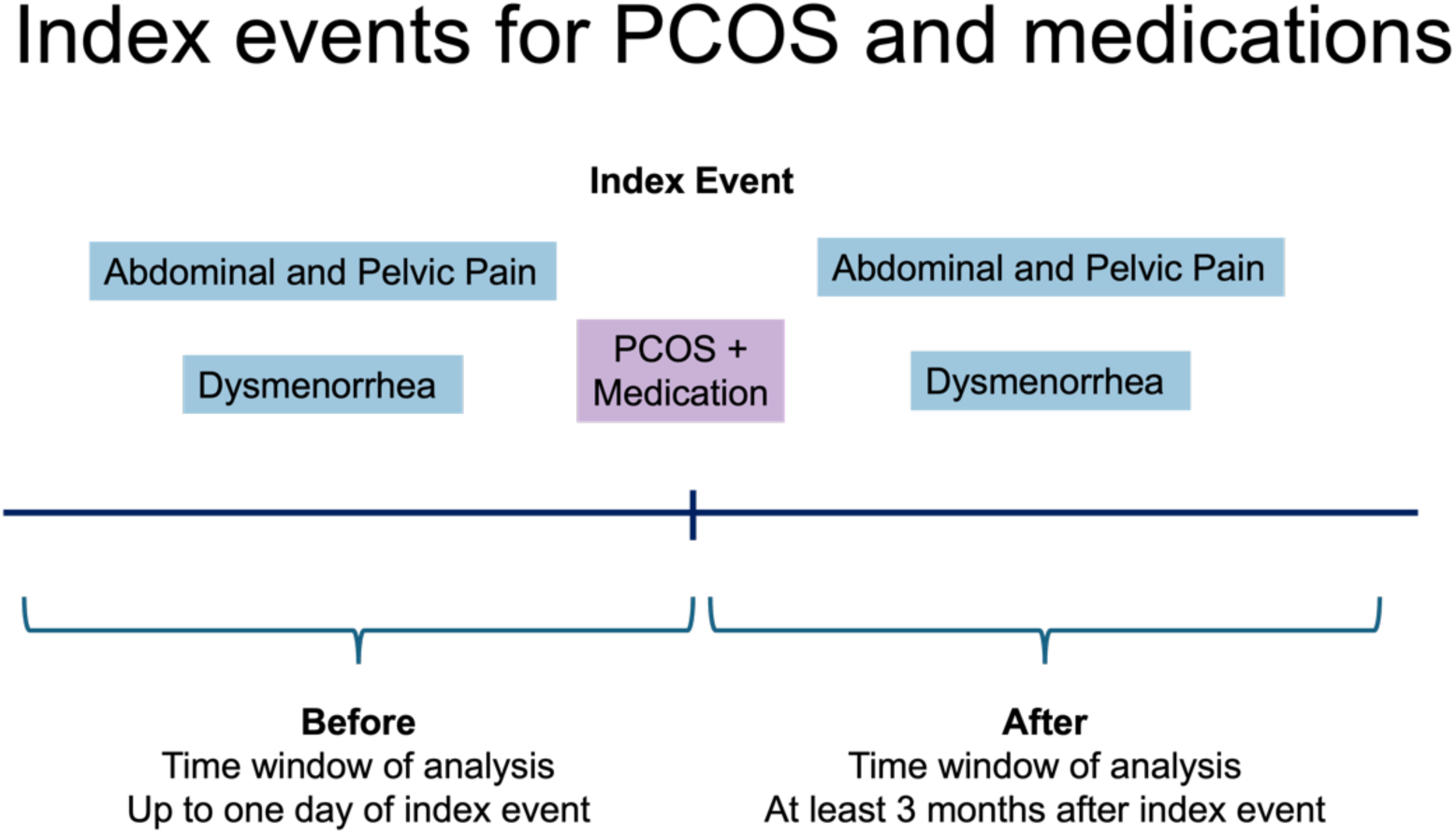
Index events for PCOS and medications. Schematic representing how the PCOS and medication events were indexed in TriNetX for both abdominal and pelvic pain and dysmenorrhea.

**Supplemental Table 2.**
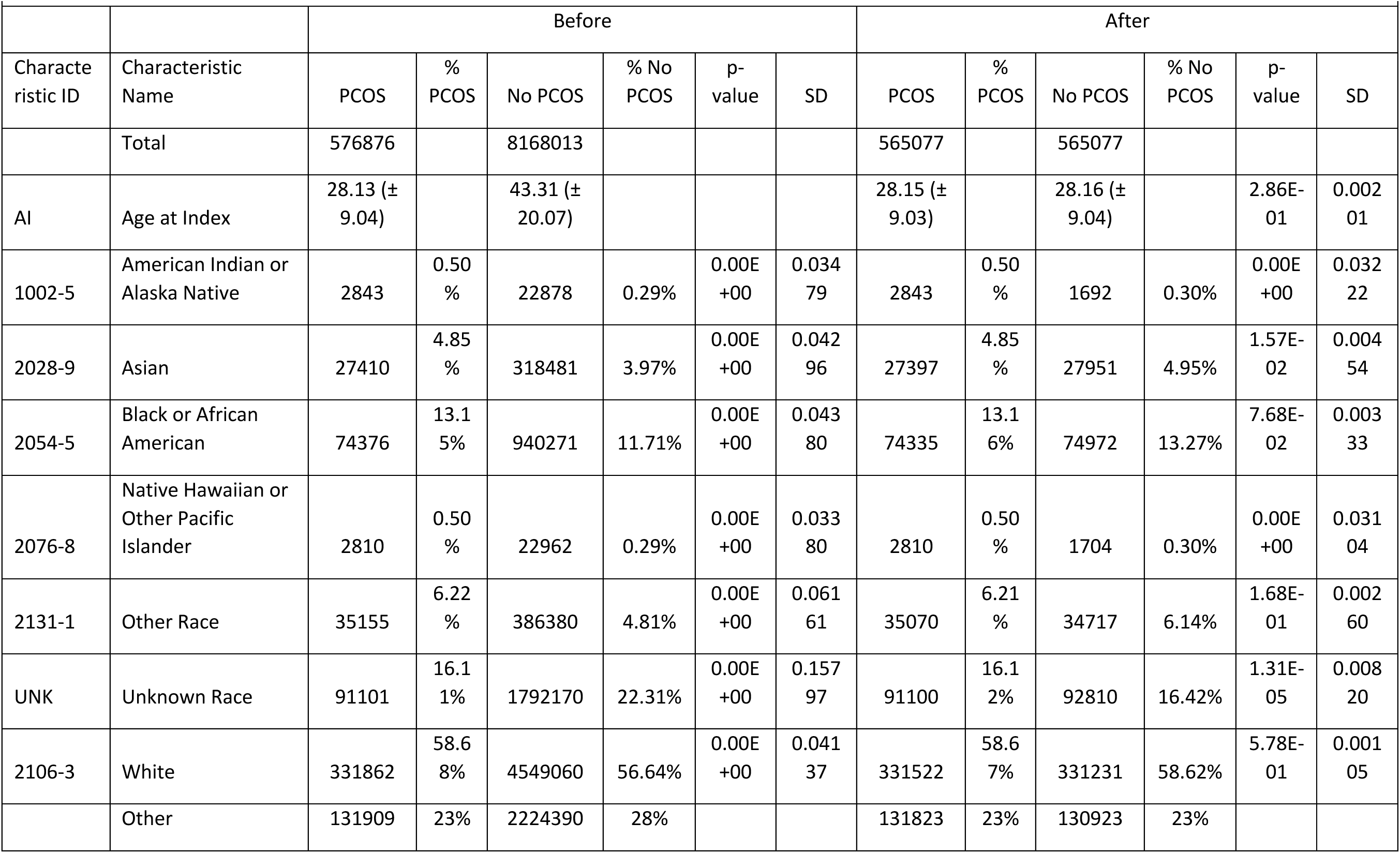
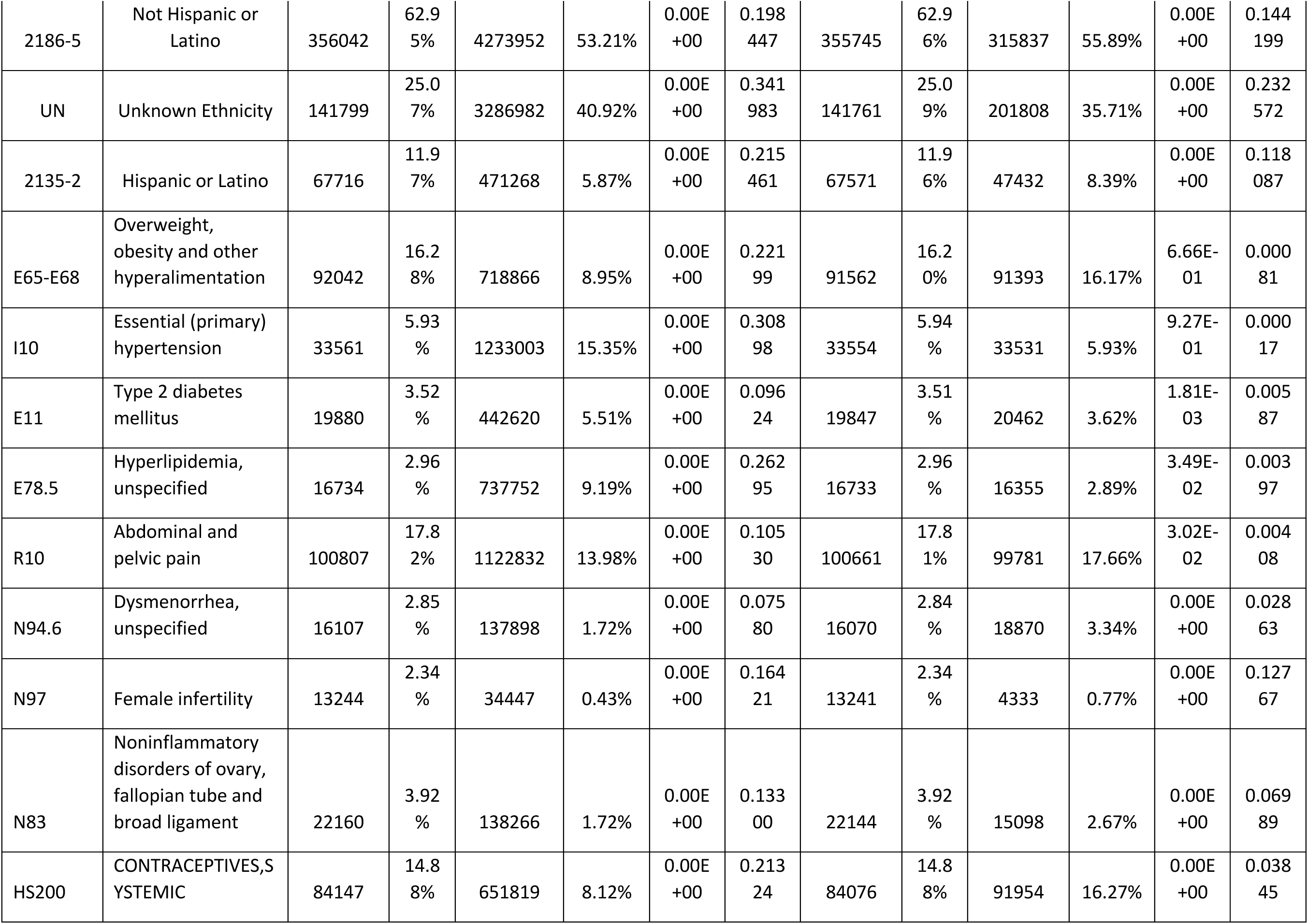
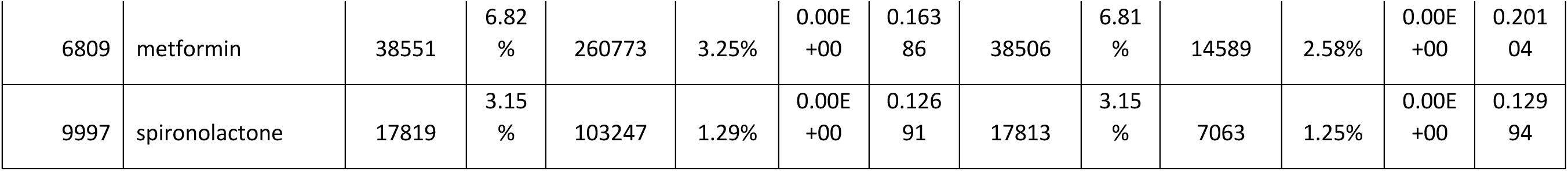
Demographic results for PCOS case and control cohorts before and after 1:1 propensity score matching. For each cohort analysis, cases were matched on the following criteria: age at the index event, self-reported race, overweight, obesity, and other hyperalimentation (ICD-10-CM E65-E69) status, type 2 diabetes mellitus (T2D) (ICD-10-CM E11) status, essential (primary) hypertension (ICD-10-CM I10) status, and hyperlipidemia, unspecified (ICD-10-CM E78.5) status. Baseline conditions were assessed up to one day before the index event.

|  |  | Before |  |  |  |  |  | After |  |  |  |  |  |
| --- | --- | --- | --- | --- | --- | --- | --- | --- | --- | --- | --- | --- | --- |
| Characteristic ID | Characteristic Name | PCOS | % PCOS | No PCOS | % No PCOS | p-value | SD | PCOS | % PCOS | No PCOS | % No PCOS | p-value | SD |
|  | Total | 576876 |  | 8168013 |  |  |  | 565077 |  | 565077 |  |  |  |
| AI | Age at Index | 28.13 (± 9.04) | | 43.31 (± 20.07) | | | | 28.15 (± 9.03) | | 28.16 (± 9.04) | | 2.86E-01 | 0.002 01 |
| 1002-5 | American Indian or Alaska Native | 2843 | 0.50 % | 22878 | 0.29% | 0.00E +00 | 0.034 79 | 2843 | 0.50 % | 1692 | 0.30% | 0.00E +00 | 0.032 22 |
| 2028-9 | Asian | 27410 | 4.85 % | 318481 | 3.97% | 0.00E +00 | 0.042 96 | 27397 | 4.85 % | 27951 | 4.95% | 1.57E-02 | 0.004 54 |
| 2054-5 | Black or African American | 74376 | 13.1 5% | 940271 | 11.71% | 0.00E +00 | 0.043 80 | 74335 | 13.1 6% | 74972 | 13.27% | 7.68E-02 | 0.003 33 |
| 2076-8 | Native Hawaiian or Other Pacific Islander | 2810 | 0.50 % | 22962 | 0.29% | 0.00E +00 | 0.033 80 | 2810 | 0.50 % | 1704 | 0.30% | 0.00E +00 | 0.031 04 |
| 2131-1 | Other Race | 35155 | 6.22 % | 386380 | 4.81% | 0.00E +00 | 0.061 61 | 35070 | 6.21 % | 34717 | 6.14% | 1.68E-01 | 0.002 60 |
| UNK | Unknown Race | 91101 | 16.1 1% | 1792170 | 22.31% | 0.00E +00 | 0.157 97 | 91100 | 16.1 2% | 92810 | 16.42% | 1.31E-05 | 0.008 20 |
| 2106-3 | White | 331862 | 58.6 8% | 4549060 | 56.64% | 0.00E +00 | 0.041 37 | 331522 | 58.6 7% | 331231 | 58.62% | 5.78E-01 | 0.001 05 |
|  | Other | 131909 | 23% | 2224390 | 28% |  |  | 131823 | 23% | 130923 | 23% |  |  |
| 2186-5 | Not Hispanic or Latino | 356042 | 62.9<br>5% | 4273952 | 53.21% | 0.00E<br>+00 | 0.198<br>447 | 355745 | 62.9<br>6% | 315837 | 55.89% | 0.00E<br>+00 | 0.144<br>199 |
| UN | Unknown Ethnicity | 141799 | 25.0<br>7% | 3286982 | 40.92% | 0.00E<br>+00 | 0.341<br>983 | 141761 | 25.0<br>9% | 201808 | 35.71% | 0.00E<br>+00 | 0.232<br>572 |
| 2135-2 | Hispanic or Latino | 67716 | 11.9<br>7% | 471268 | 5.87% | 0.00E<br>+00 | 0.215<br>461 | 67571 | 11.9<br>6% | 47432 | 8.39% | 0.00E<br>+00 | 0.118<br>087 |
| E65-E68 | Overweight, obesity and other hyperalimentation | 92042 | 16.2<br>8% | 718866 | 8.95% | 0.00E<br>+00 | 0.221<br>99 | 91562 | 16.2<br>0% | 91393 | 16.17% | 6.66E-01 | 0.000<br>81 |
| I10 | Essential (primary) hypertension | 33561 | 5.93<br>% | 1233003 | 15.35% | 0.00E<br>+00 | 0.308<br>98 | 33554 | 5.94<br>% | 33531 | 5.93% | 9.27E-01 | 0.000<br>17 |
| E11 | Type 2 diabetes mellitus | 19880 | 3.52<br>% | 442620 | 5.51% | 0.00E<br>+00 | 0.096<br>24 | 19847 | 3.51<br>% | 20462 | 3.62% | 1.81E-03 | 0.005<br>87 |
| E78.5 | Hyperlipidemia, unspecified | 16734 | 2.96<br>% | 737752 | 9.19% | 0.00E<br>+00 | 0.262<br>95 | 16733 | 2.96<br>% | 16355 | 2.89% | 3.49E-02 | 0.003<br>97 |
| R10 | Abdominal and pelvic pain | 100807 | 17.8<br>2% | 1122832 | 13.98% | 0.00E<br>+00 | 0.105<br>30 | 100661 | 17.8<br>1% | 99781 | 17.66% | 3.02E-02 | 0.004<br>08 |
| N94.6 | Dysmenorrhea, unspecified | 16107 | 2.85<br>% | 137898 | 1.72% | 0.00E<br>+00 | 0.075<br>80 | 16070 | 2.84<br>% | 18870 | 3.34% | 0.00E<br>+00 | 0.028<br>63 |
| N97 | Female infertility | 13244 | 2.34<br>% | 34447 | 0.43% | 0.00E<br>+00 | 0.164<br>21 | 13241 | 2.34<br>% | 4333 | 0.77% | 0.00E<br>+00 | 0.127<br>67 |
| N83 | Noninflammatory disorders of ovary, fallopian tube and broad ligament | 22160 | 3.92<br>% | 138266 | 1.72% | 0.00E<br>+00 | 0.133<br>00 | 22144 | 3.92<br>% | 15098 | 2.67% | 0.00E<br>+00 | 0.069<br>89 |
| HS200 | CONTRACEPTIVES,SYSTEMIC | 84147 | 14.8<br>8% | 651819 | 8.12% | 0.00E<br>+00 | 0.213<br>24 | 84076 | 14.8<br>8% | 91954 | 16.27% | 0.00E<br>+00 | 0.038<br>45 |
| 6809 | metformin | 38551 | 6.82<br>% | 260773 | 3.25% | 0.00E<br>+00 | 0.163<br>86 | 38506 | 6.81<br>% | 14589 | 2.58% | 0.00E<br>+00 | 0.201<br>04 |
| 9997 | spironolactone | 17819 | 3.15<br>% | 103247 | 1.29% | 0.00E<br>+00 | 0.126<br>91 | 17813 | 3.15<br>% | 7063 | 1.25% | 0.00E<br>+00 | 0.129<br>94 |

**Table 3 - Figure Supplement 6.**
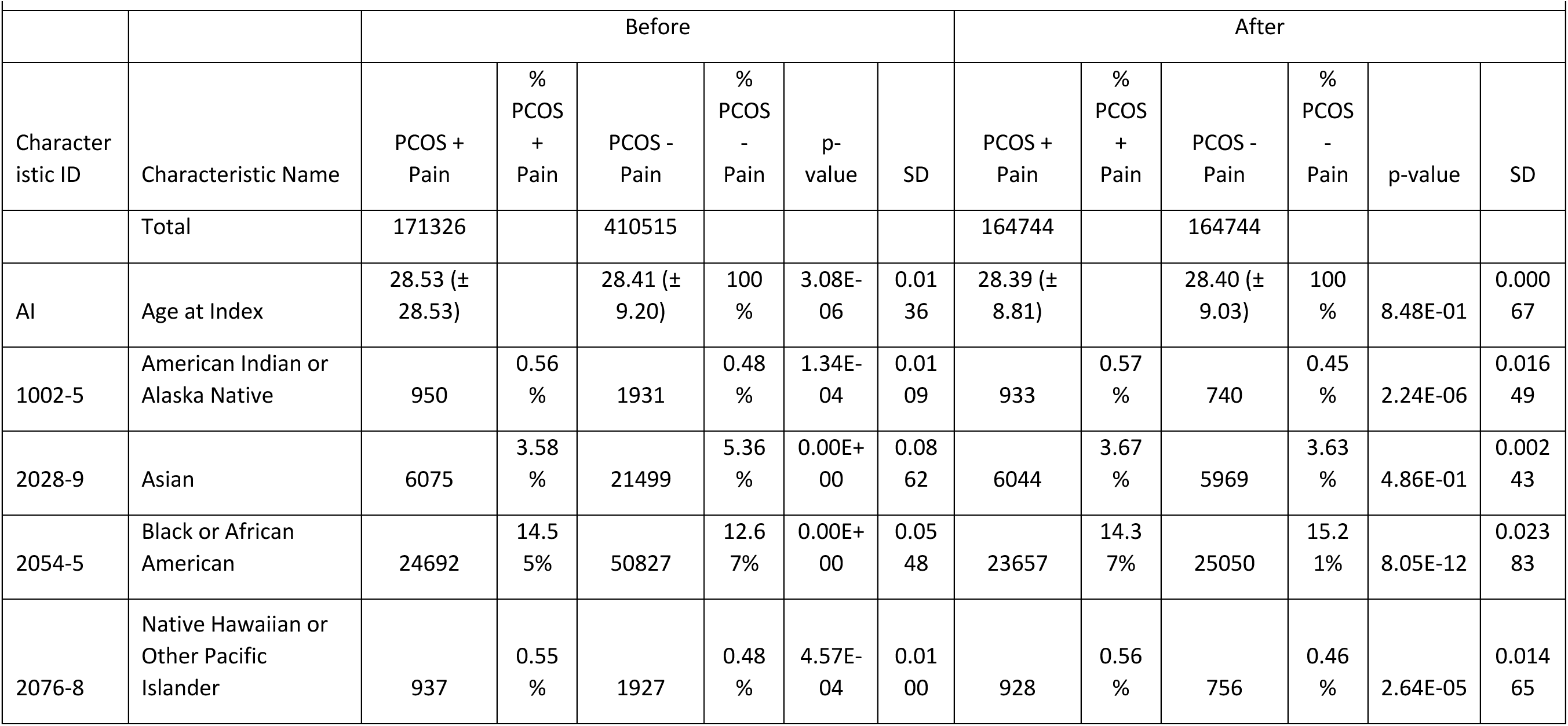
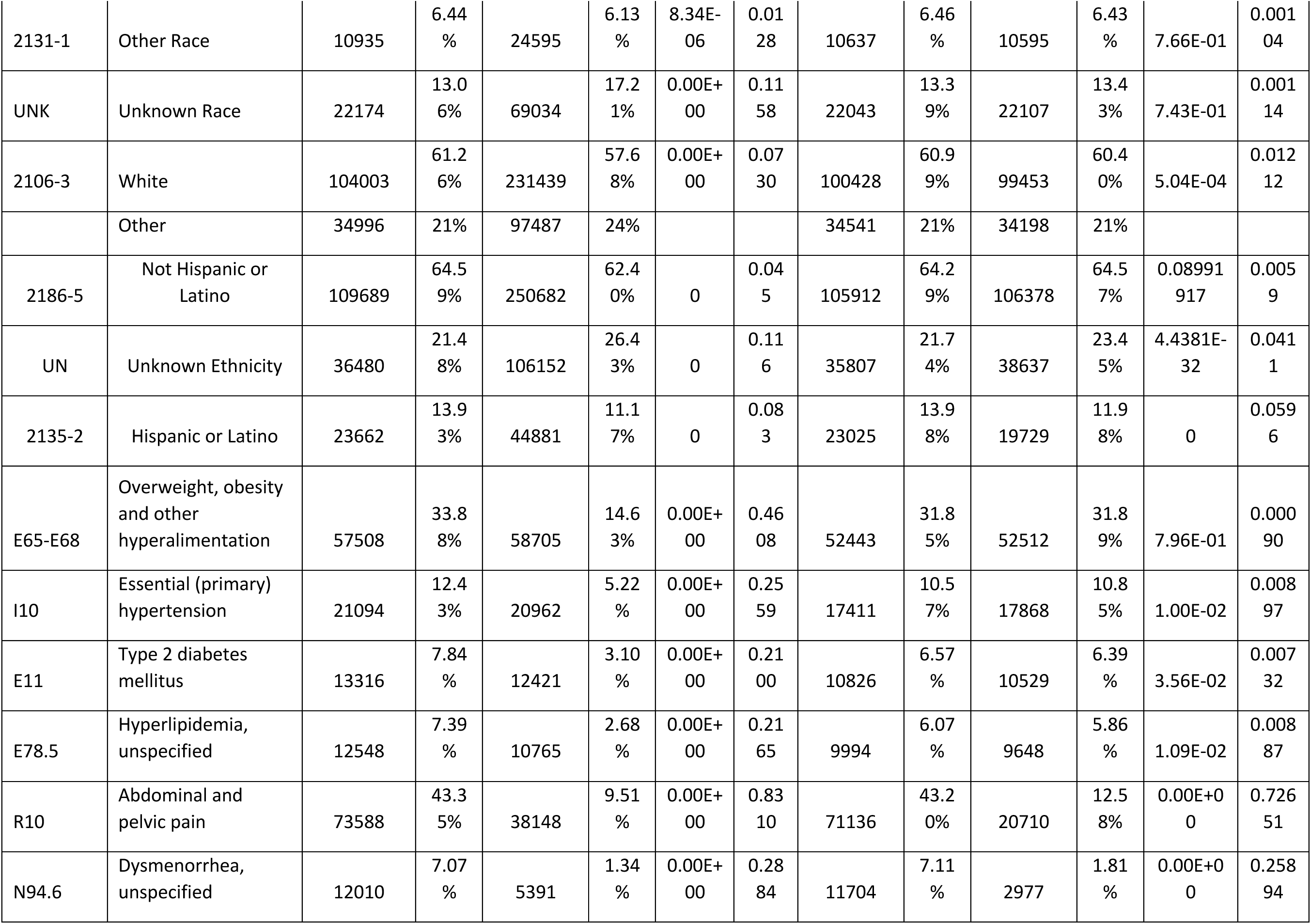
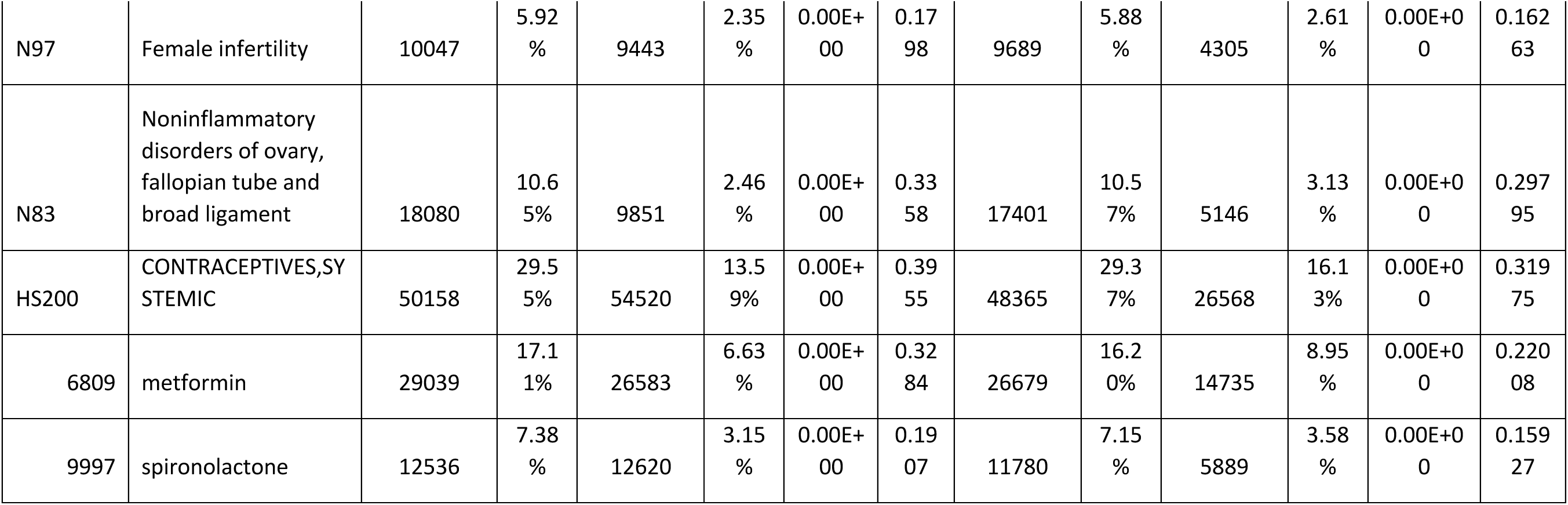
Demographic and baseline characteristics for PCOS and Pain case and control cohorts before and after 1:1 propensity score matching. For each cohort analysis, cases were matched on the following criteria: age at the index event, self-reported race, overweight, obesity, and other hyperalimentation (ICD-10-CM E65-E69) status, type 2 diabetes mellitus (T2D) (ICD-10-CM E11) status, essential (primary) hypertension (ICD-10-CM I10) status, and hyperlipidemia, unspecified (ICD-10-CM E78.5) status. Baseline conditions were assessed up to one day before the index event.

|  |  |  |  |  |  |  |  |  |  |  |  |  |  |
| --- | --- | --- | --- | --- | --- | --- | --- | --- | --- | --- | --- | --- | --- |
|  |  | Before |  |  |  |  |  | After |  |  |  |  |  |
| Character<br>istic ID | Characteristic Name | PCOS +<br>Pain | %<br>PCOS<br>+<br>Pain | PCOS -<br>Pain | %<br>PCOS<br>-<br>Pain | p-value | SD | PCOS +<br>Pain | %<br>PCOS<br>+<br>Pain | PCOS -<br>Pain | %<br>PCOS<br>-<br>Pain | p-value | SD |
|  | Total | 171326 |  | 410515 |  |  |  | 164744 |  | 164744 |  |  |  |
| AI | Age at Index | 28.53 (±<br>28.53) |  | 28.41 (±<br>9.20) | 100<br>% | 3.08E-06 | 0.01<br>36 | 28.39 (±<br>8.81) |  | 28.40 (±<br>9.03) | 100<br>% | 8.48E-01 | 0.000<br>67 |
| 1002-5 | American Indian or<br>Alaska Native | 950 | 0.56<br>% | 1931 | 0.48<br>% | 1.34E-04 | 0.01<br>09 | 933 | 0.57<br>% | 740 | 0.45<br>% | 2.24E-06 | 0.016<br>49 |
| 2028-9 | Asian | 6075 | 3.58<br>% | 21499 | 5.36<br>% | 0.00E+<br>00 | 0.08<br>62 | 6044 | 3.67<br>% | 5969 | 3.63<br>% | 4.86E-01 | 0.002<br>43 |
| 2054-5 | Black or African<br>American | 24692 | 14.5<br>5% | 50827 | 12.6<br>7% | 0.00E+<br>00 | 0.05<br>48 | 23657 | 14.3<br>7% | 25050 | 15.2<br>1% | 8.05E-12 | 0.023<br>83 |
| 2076-8 | Native Hawaiian or<br>Other Pacific<br>Islander | 937 | 0.55<br>% | 1927 | 0.48<br>% | 4.57E-04 | 0.01<br>00 | 928 | 0.56<br>% | 756 | 0.46<br>% | 2.64E-05 | 0.014<br>65 |
| 2131-1 | Other Race | 10935 | 6.44<br>% | 24595 | 6.13<br>% | 8.34E-06 | 0.01<br>28 | 10637 | 6.46<br>% | 10595 | 6.43<br>% | 7.66E-01 | 0.001<br>04 |
| UNK | Unknown Race | 22174 | 13.0<br>6% | 69034 | 17.2<br>1% | 0.00E+00 | 0.11<br>58 | 22043 | 13.3<br>9% | 22107 | 13.4<br>3% | 7.43E-01 | 0.001<br>14 |
| 2106-3 | White | 104003 | 61.2<br>6% | 231439 | 57.6<br>8% | 0.00E+00 | 0.07<br>30 | 100428 | 60.9<br>9% | 99453 | 60.4<br>0% | 5.04E-04 | 0.012<br>12 |
|  | Other | 34996 | 21% | 97487 | 24% |  |  | 34541 | 21% | 34198 | 21% |  |  |
| 2186-5 | Not Hispanic or Latino | 109689 | 64.5<br>9% | 250682 | 62.4<br>0% | 0 | 0.04<br>5 | 105912 | 64.2<br>9% | 106378 | 64.5<br>7% | 0.08991<br>917 | 0.005<br>9 |
| UN | Unknown Ethnicity | 36480 | 21.4<br>8% | 106152 | 26.4<br>3% | 0 | 0.11<br>6 | 35807 | 21.7<br>4% | 38637 | 23.4<br>5% | 4.4381E-32 | 0.041<br>1 |
| 2135-2 | Hispanic or Latino | 23662 | 13.9<br>3% | 44881 | 11.1<br>7% | 0 | 0.08<br>3 | 23025 | 13.9<br>8% | 19729 | 11.9<br>8% | 0 | 0.059<br>6 |
| E65-E68 | Overweight, obesity and other hyperalimentation | 57508 | 33.8<br>8% | 58705 | 14.6<br>3% | 0.00E+00 | 0.46<br>08 | 52443 | 31.8<br>5% | 52512 | 31.8<br>9% | 7.96E-01 | 0.000<br>90 |
| I10 | Essential (primary) hypertension | 21094 | 12.4<br>3% | 20962 | 5.22<br>% | 0.00E+00 | 0.25<br>59 | 17411 | 10.5<br>7% | 17868 | 10.8<br>5% | 1.00E-02 | 0.008<br>97 |
| E11 | Type 2 diabetes mellitus | 13316 | 7.84<br>% | 12421 | 3.10<br>% | 0.00E+00 | 0.21<br>00 | 10826 | 6.57<br>% | 10529 | 6.39<br>% | 3.56E-02 | 0.007<br>32 |
| E78.5 | Hyperlipidemia, unspecified | 12548 | 7.39<br>% | 10765 | 2.68<br>% | 0.00E+00 | 0.21<br>65 | 9994 | 6.07<br>% | 9648 | 5.86<br>% | 1.09E-02 | 0.008<br>87 |
| R10 | Abdominal and pelvic pain | 73588 | 43.3<br>5% | 38148 | 9.51<br>% | 0.00E+00 | 0.83<br>10 | 71136 | 43.2<br>0% | 20710 | 12.5<br>8% | 0.00E+0<br>0 | 0.726<br>51 |
| N94.6 | Dysmenorrhea, unspecified | 12010 | 7.07<br>% | 5391 | 1.34<br>% | 0.00E+00 | 0.28<br>84 | 11704 | 7.11<br>% | 2977 | 1.81<br>% | 0.00E+0<br>0 | 0.258<br>94 |
| N97 | Female infertility | 10047 | 5.92<br>% | 9443 | 2.35<br>% | 0.00E+<br>00 | 0.17<br>98 | 9689 | 5.88<br>% | 4305 | 2.61<br>% | 0.00E+0<br>0 | 0.162<br>63 |
| N83 | Noninflammatory disorders of ovary, fallopian tube and broad ligament | 18080 | 10.6<br>5% | 9851 | 2.46<br>% | 0.00E+<br>00 | 0.33<br>58 | 17401 | 10.5<br>7% | 5146 | 3.13<br>% | 0.00E+0<br>0 | 0.297<br>95 |
| HS200 | CONTRACEPTIVES,SY<br>STEMIC | 50158 | 29.5<br>5% | 54520 | 13.5<br>9% | 0.00E+<br>00 | 0.39<br>55 | 48365 | 29.3<br>7% | 26568 | 16.1<br>3% | 0.00E+0<br>0 | 0.319<br>75 |
| 6809 | metformin | 29039 | 17.1<br>1% | 26583 | 6.63<br>% | 0.00E+<br>00 | 0.32<br>84 | 26679 | 16.2<br>0% | 14735 | 8.95<br>% | 0.00E+0<br>0 | 0.220<br>08 |
| 9997 | spironolactone | 12536 | 7.38<br>% | 12620 | 3.15<br>% | 0.00E+<br>00 | 0.19<br>07 | 11780 | 7.15<br>% | 5889 | 3.58<br>% | 0.00E+0<br>0 | 0.159<br>27 |

**Figure 4 – Figure Supplement 7.**
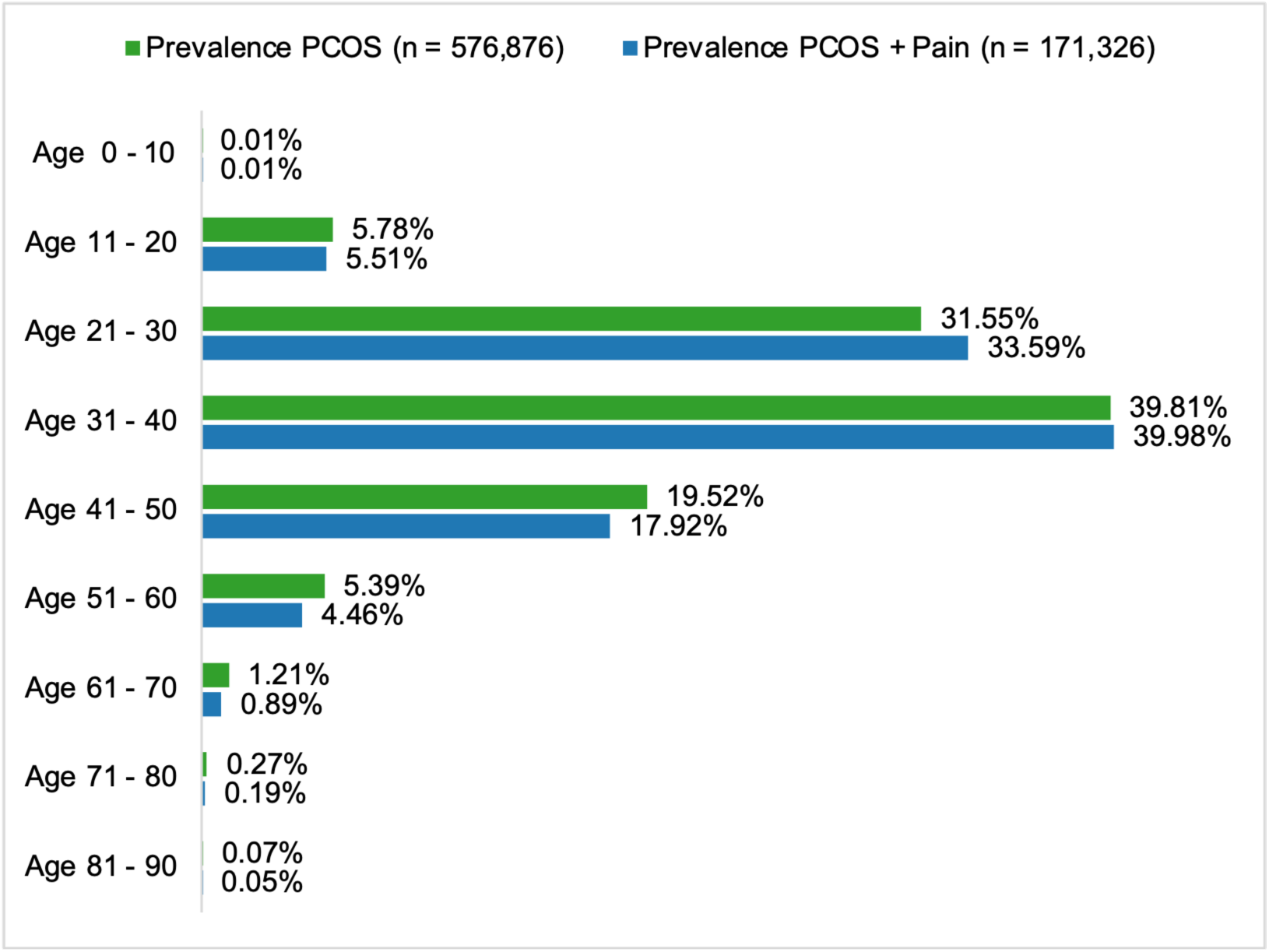
Age-stratified Prevalence of PCOS and PCOS and Pain. Bar plots show the prevalence (%) of overall PCOS (green) and PCOS and Pain (blue) stratified by 10-year age groups. The total number of women with PCOS is 576,876 and the total number of women with PCOS and Pain is 171,326.

**Supplemental Table 4 – Figure Supplement 8.** Counts and prevalence (%) of PCOS cases and controls for self-reported race groups.

|  |  |  |  |
| --- | --- | --- | --- |
| Self-reported race group | PCOS | No PCOS | Prevalence (%) |
| Asian | 4810 | 16345 | 22.74% |
| Black or African American | 16887 | 35703 | 32.11% |
| Other | 25909 | 78786 | 24.75% |
| White | 87077 | 196103 | 30.75% |

**Supplemental Table 5 - Figure Supplement 9.**
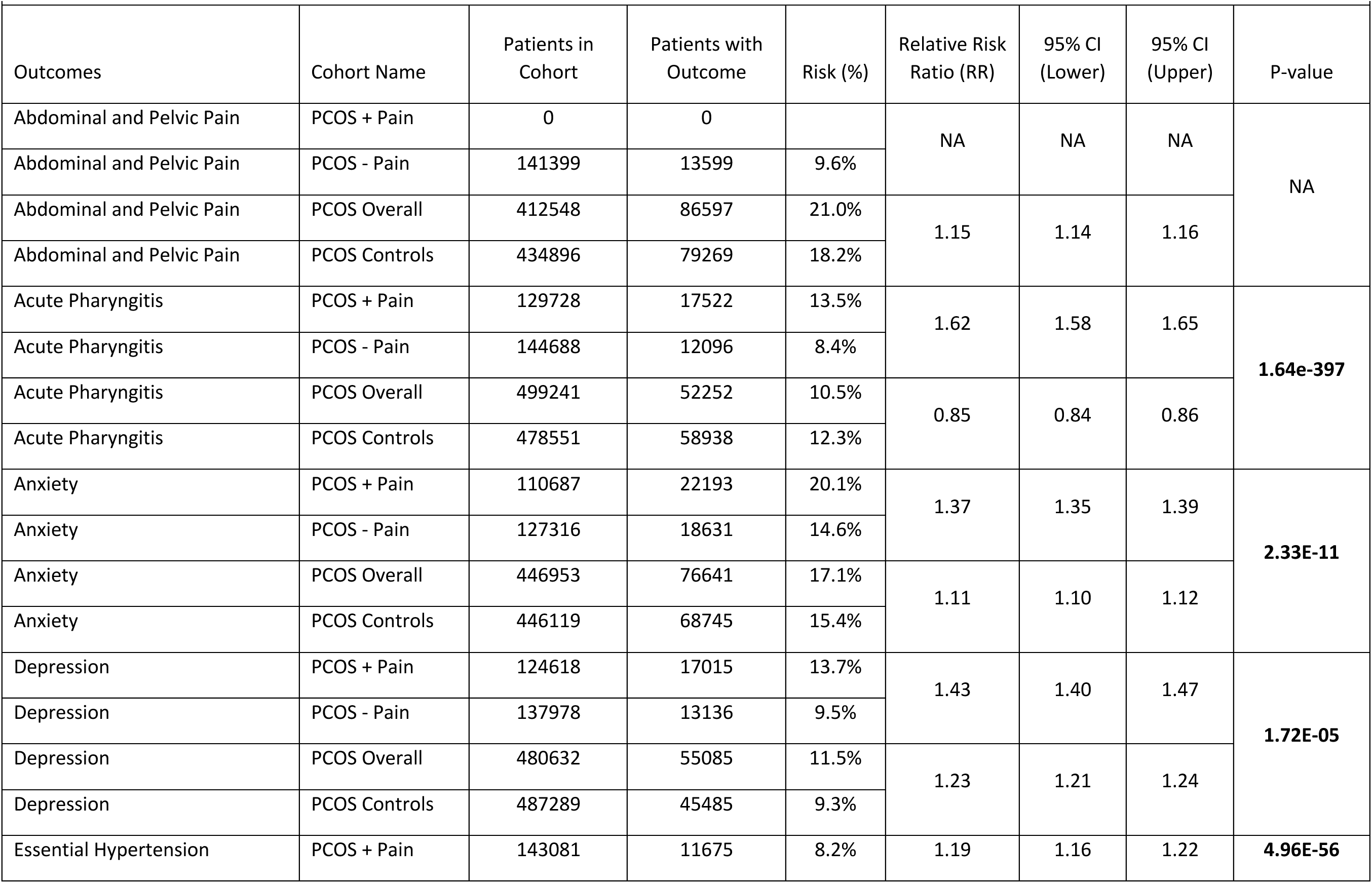
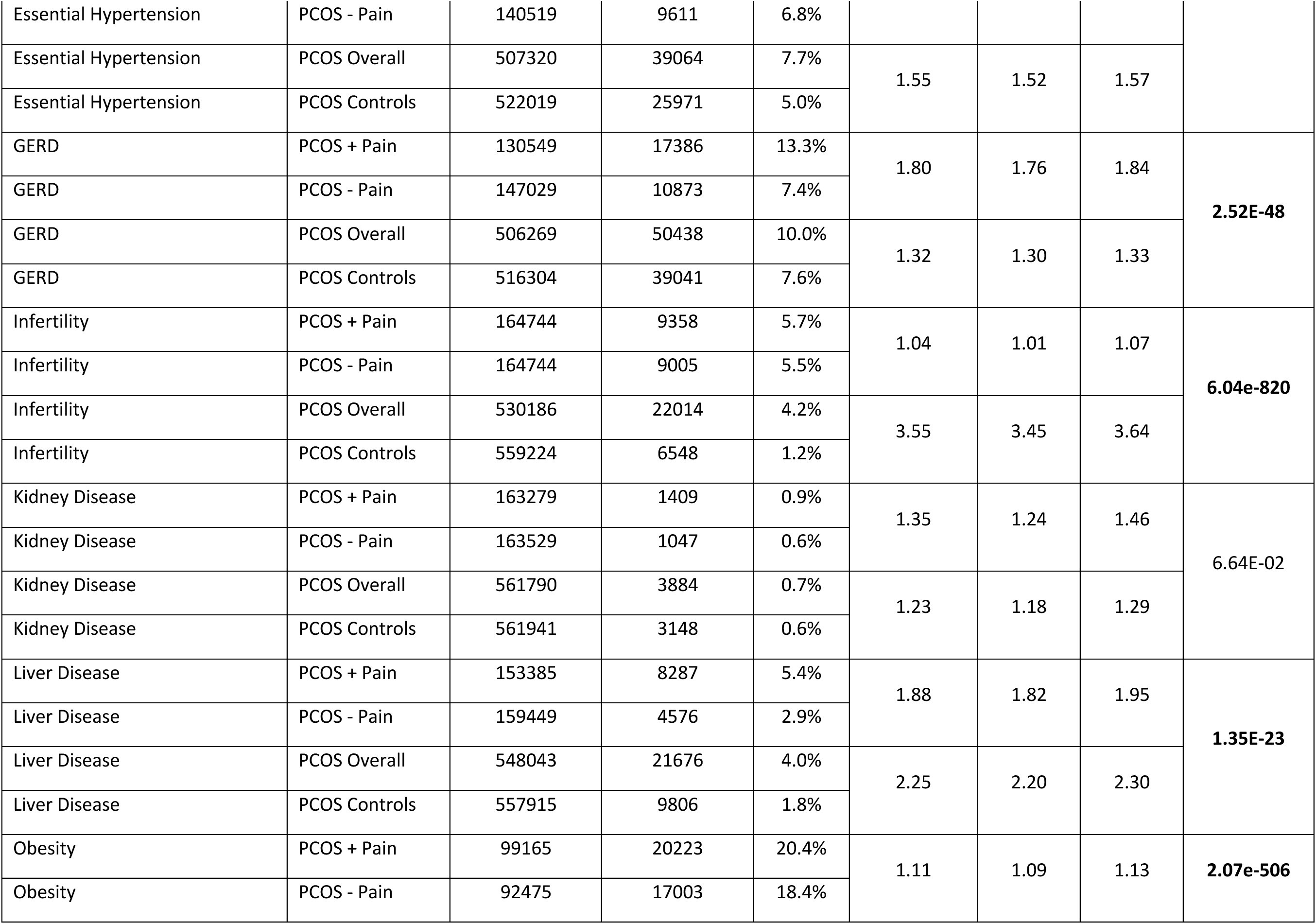
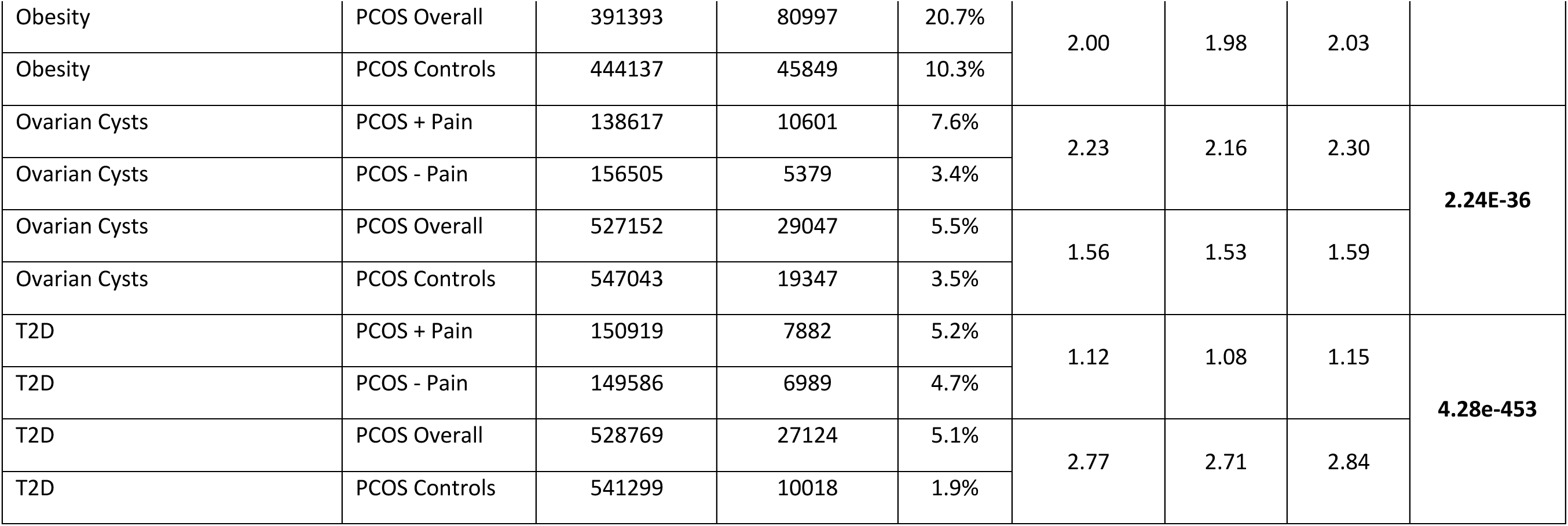
Relative risk ratios (RR) for PCOS and PCOS and Pain cases and control cohorts. Significant differences in RR between PCOS and PCOS and Pain cohorts are bolded.

**Supplemental Table 6 - Figure Supplement 10.**
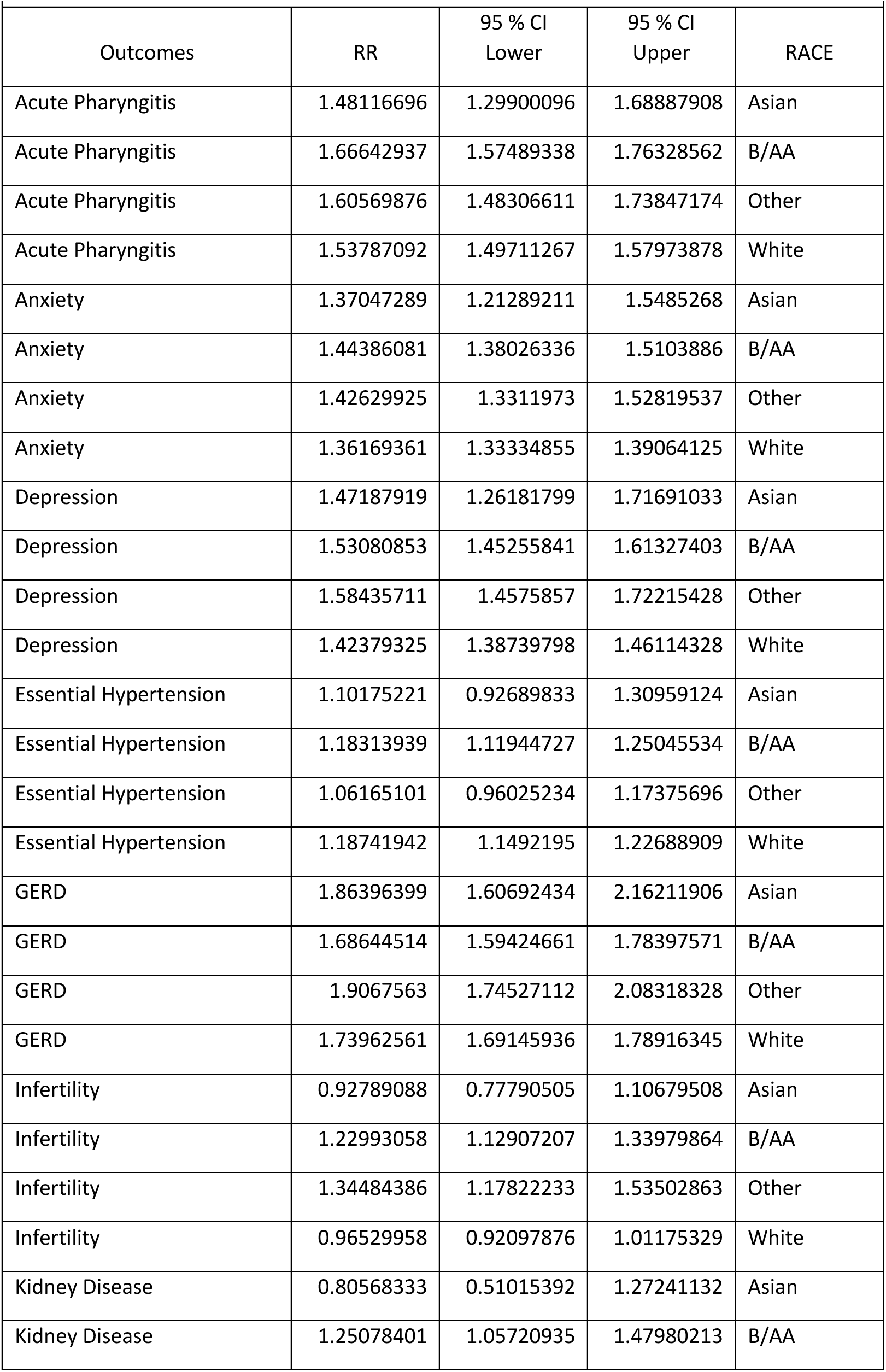
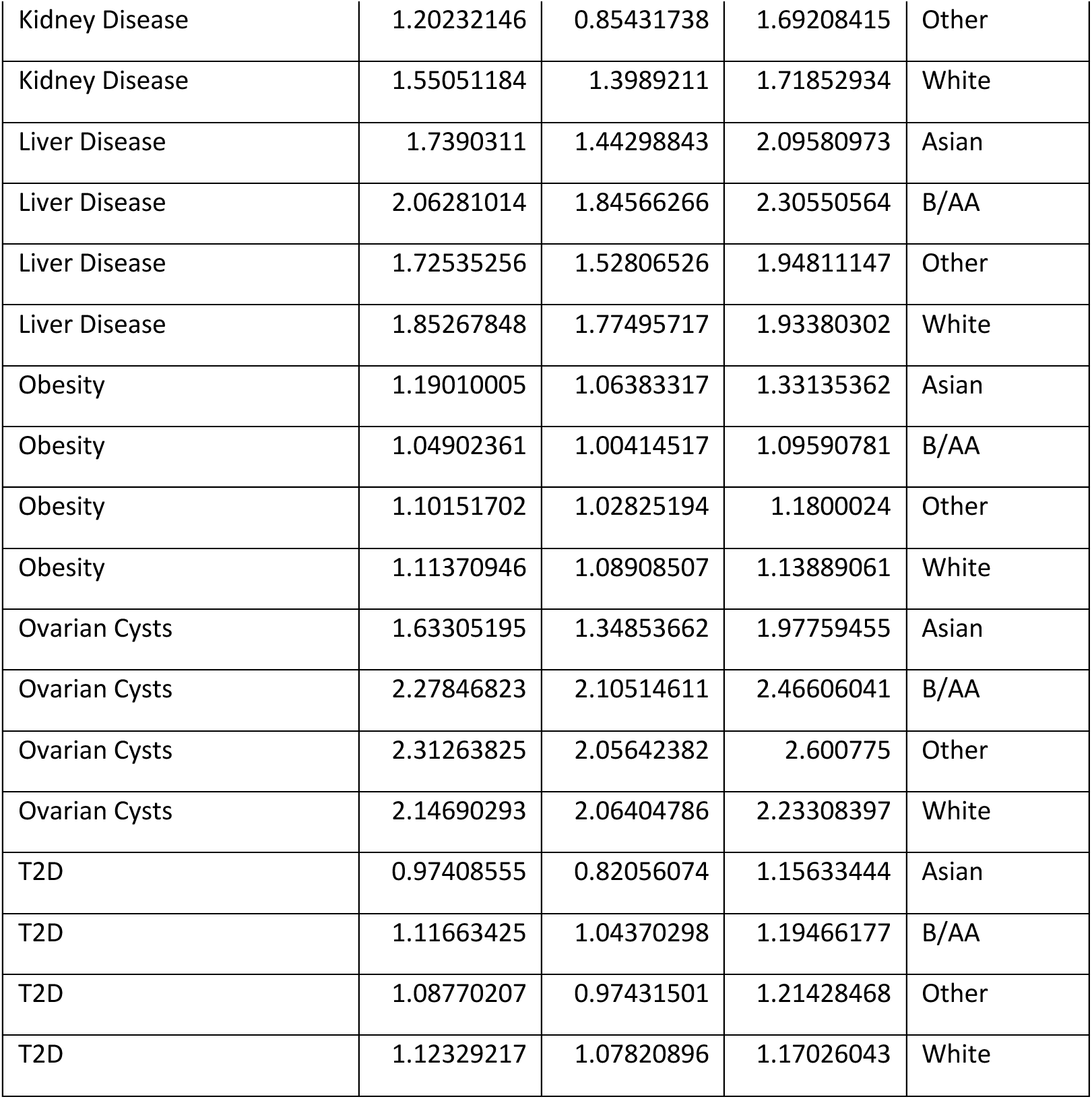
Relative risk ratios (RR) for race-stratified PCOS and Pain cohorts.

|  |  |  |  |  |
| --- | --- | --- | --- | --- |
| Outcomes | RR | 95 % CI<br>Lower | 95 % CI<br>Upper | RACE |
| Acute Pharyngitis | 1.48116696 | 1.29900096 | 1.68887908 | Asian |
| Acute Pharyngitis | 1.66642937 | 1.57489338 | 1.76328562 | B/AA |
| Acute Pharyngitis | 1.60569876 | 1.48306611 | 1.73847174 | Other |
| Acute Pharyngitis | 1.53787092 | 1.49711267 | 1.57973878 | White |
| Anxiety | 1.37047289 | 1.21289211 | 1.5485268 | Asian |
| Anxiety | 1.44386081 | 1.38026336 | 1.5103886 | B/AA |
| Anxiety | 1.42629925 | 1.3311973 | 1.52819537 | Other |
| Anxiety | 1.36169361 | 1.33334855 | 1.39064125 | White |
| Depression | 1.47187919 | 1.26181799 | 1.71691033 | Asian |
| Depression | 1.53080853 | 1.45255841 | 1.61327403 | B/AA |
| Depression | 1.58435711 | 1.4575857 | 1.72215428 | Other |
| Depression | 1.42379325 | 1.38739798 | 1.46114328 | White |
| Essential Hypertension | 1.10175221 | 0.92689833 | 1.30959124 | Asian |
| Essential Hypertension | 1.18313939 | 1.11944727 | 1.25045534 | B/AA |
| Essential Hypertension | 1.06165101 | 0.96025234 | 1.17375696 | Other |
| Essential Hypertension | 1.18741942 | 1.1492195 | 1.22688909 | White |
| GERD | 1.86396399 | 1.60692434 | 2.16211906 | Asian |
| GERD | 1.68644514 | 1.59424661 | 1.78397571 | B/AA |
| GERD | 1.9067563 | 1.74527112 | 2.08318328 | Other |
| GERD | 1.73962561 | 1.69145936 | 1.78916345 | White |
| Infertility | 0.92789088 | 0.77790505 | 1.10679508 | Asian |
| Infertility | 1.22993058 | 1.12907207 | 1.33979864 | B/AA |
| Infertility | 1.34484386 | 1.17822233 | 1.53502863 | Other |
| Infertility | 0.96529958 | 0.92097876 | 1.01175329 | White |
| Kidney Disease | 0.80568333 | 0.51015392 | 1.27241132 | Asian |
| Kidney Disease | 1.25078401 | 1.05720935 | 1.47980213 | B/AA |
| Kidney Disease | 1.20232146 | 0.85431738 | 1.69208415 | Other |
| Kidney Disease | 1.55051184 | 1.3989211 | 1.71852934 | White |
| Liver Disease | 1.7390311 | 1.44298843 | 2.09580973 | Asian |
| Liver Disease | 2.06281014 | 1.84566266 | 2.30550564 | B/AA |
| Liver Disease | 1.72535256 | 1.52806526 | 1.94811147 | Other |
| Liver Disease | 1.85267848 | 1.77495717 | 1.93380302 | White |
| Obesity | 1.19010005 | 1.06383317 | 1.33135362 | Asian |
| Obesity | 1.04902361 | 1.00414517 | 1.09590781 | B/AA |
| Obesity | 1.10151702 | 1.02825194 | 1.1800024 | Other |
| Obesity | 1.11370946 | 1.08908507 | 1.13889061 | White |
| Ovarian Cysts | 1.63305195 | 1.34853662 | 1.97759455 | Asian |
| Ovarian Cysts | 2.27846823 | 2.10514611 | 2.46606041 | B/AA |
| Ovarian Cysts | 2.31263825 | 2.05642382 | 2.600775 | Other |
| Ovarian Cysts | 2.14690293 | 2.06404786 | 2.23308397 | White |
| T2D | 0.97408555 | 0.82056074 | 1.15633444 | Asian |
| T2D | 1.11663425 | 1.04370298 | 1.19466177 | B/AA |
| T2D | 1.08770207 | 0.97431501 | 1.21428468 | Other |
| T2D | 1.12329217 | 1.07820896 | 1.17026043 | White |

**Supplemental Table 7 - Figure Supplement 11.** Significance for relative risk ratios for each self-reported race combination. Both P-value (P) and adjusted p-values using Benjamini-Hochberg (P(BH)) are reported. Significant differences in RR between self-reported race groups are bolded.

| Future Health Outcome | Group 1 | Group 2 | Z | P | P(BH) |
| --- | --- | --- | --- | --- | --- |
| Acute Pharyngitis | Asian | B/AA | -1.6166987 | 0.10594334 | 0.31783001 |
| Acute Pharyngitis | Asian | Other | -1.0314123 | 0.3023475 | 0.46968711 |
| Acute Pharyngitis | Asian | White | -0.5496948 | 0.58252871 | 0.58252871 |
| Acute Pharyngitis | B/AA | Other | 0.74639393 | 0.45542949 | 0.54651538 |
| Acute Pharyngitis | B/AA | White | 2.5154627 | 0.01188763 | 0.07132575 |
| Acute Pharyngitis | Other | White | 1.00868706 | 0.31312474 | 0.46968711 |
| Anxiety | Asian | B/AA | -0.7853388 | 0.43225496 | 0.86450992 |
| Anxiety | Asian | Other | -0.5578196 | 0.57696755 | 0.86545132 |
| Anxiety | Asian | White | 0.10162617 | 0.91905341 | 0.91905341 |
| Anxiety | B/AA | Other | 0.29106415 | 0.77100226 | 0.91905341 |
| Anxiety | B/AA | White | 2.30990701 | 0.0208933 | 0.12535982 |
| Anxiety | Other | White | 1.25941664 | 0.20787988 | 0.62363965 |
| Depression | Asian | B/AA | -0.4729643 | 0.63623868 | 0.676734 |
| Depression | Asian | Other | -0.8241903 | 0.40983142 | 0.676734 |
| Depression | Asian | White | 0.41692399 | 0.676734 | 0.676734 |
| Depression | B/AA | Other | -0.6839542 | 0.49400409 | 0.676734 |
| Depression | B/AA | White | 2.42762719 | 0.01519796 | **0.04940984** |
| Depression | Other | White | 2.39833165 | 0.01646995 | **0.04940984** |
| Essential Hypertension | Asian | B/AA | -0.7698166 | 0.44140868 | 0.66211302 |
| Essential Hypertension | Asian | Other | 0.36361672 | 0.71614424 | 0.85937309 |
| Essential Hypertension | Asian | White | -0.8344677 | 0.4040175 | 0.66211302 |
| Essential Hypertension | B/AA | Other | 1.8526185 | 0.06393706 | 0.19181118 |
| Essential Hypertension | B/AA | White | -0.1101123 | 0.91232028 | 0.91232028 |
| Essential Hypertension | Other | White | -2.0784656 | 0.0376665 | 0.19181118 |
| GERD | Asian | B/AA | 1.23623073 | 0.21637281 | 0.43274562 |
| GERD | Asian | Other | -0.2575035 | 0.79679012 | 0.79679012 |
| GERD | Asian | White | 0.89599097 | 0.37025759 | 0.4443091 |
| GERD | B/AA | Other | -2.2953454 | 0.02171334 | 0.13028004 |
| GERD | B/AA | White | -0.9683209 | 0.33288412 | 0.4443091 |
| GERD | Other | White | 1.93661501 | 0.05279242 | 0.15837727 |
| Infertility | Asian | B/AA | -2.8183597 | 0.00482697 | **0.00724046** |
| Infertility | Asian | Other | -3.3001771 | 0.00096624 | **0.00193248** |
| Infertility | Asian | White | -0.4245574 | 0.67115934 | 0.67115934 |
| Infertility | B/AA | Other | -1.111312 | 0.26643407 | 0.31972089 |
| Infertility | B/AA | White | 4.86428327 | 1.15E-06 | **6.89E-06** |
| Infertility | Other | White | 4.62996514 | 3.66E-06 | **1.10E-05** |
| Kidney Disease | Asian | B/AA | -1.7704376 | 0.07665427 | 0.15330853 |
| Kidney Disease | Asian | Other | -1.3750696 | 0.16910986 | 0.20293183 |
| Kidney Disease | Asian | White | -2.7392558 | 0.00615784 | **0.03694707** |
| Kidney Disease | B/AA | Other | 0.2033752 | 0.83884178 | 0.83884178 |
| Kidney Disease | B/AA | White | -2.1359645 | 0.03268231 | 0.09804693 |
| Kidney Disease | Other | White | -1.3968767 | 0.16245065 | 0.20293183 |
| Liver Disease | Asian | B/AA | -1.5404237 | 0.12345711 | 0.24691422 |
| Liver Disease | Asian | Other | 0.06951799 | 0.94457731 | 0.94457731 |
| Liver Disease | Asian | White | -0.6480213 | 0.51697119 | 0.62036542 |
| Liver Disease | B/AA | Other | 2.12621006 | 0.03348577 | 0.20091464 |
| Liver Disease | B/AA | White | 1.76655909 | 0.0773021 | 0.23190629 |
| Liver Disease | Other | White | -1.083748 | 0.27847656 | 0.41771484 |
| Obesity | Asian | B/AA | 2.05439618 | 0.03993736 | 0.11981209 |
| Obesity | Asian | Other | 1.15206197 | 0.24929561 | 0.30666863 |
| Obesity | Asian | White | 1.13695477 | 0.25555719 | 0.30666863 |
| Obesity | B/AA | Other | -1.1736814 | 0.24052266 | 0.30666863 |
| Obesity | B/AA | White | -2.3881842 | 0.01693185 | 0.10159113 |
| Obesity | Other | White | -0.2981351 | 0.76560009 | 0.76560009 |
| Ovarian Cysts | Asian | B/AA | -3.1514691 | 0.00162451 | **0.00717584** |
| Ovarian Cysts | Asian | Other | -3.0366859 | 0.00239195 | **0.00717584** |
| Ovarian Cysts | Asian | White | -2.7436718 | 0.00607562 | **0.01215125** |
| Ovarian Cysts | B/AA | Other | -0.2060601 | 0.83674398 | 0.83674398 |
| Ovarian Cysts | B/AA | White | 1.31921453 | 0.1870974 | 0.2806461 |
| Ovarian Cysts | Other | White | 1.17691992 | 0.23922747 | 0.28707296 |
| T2D | Asian | B/AA | -1.4522022 | 0.1464454 | 0.43933621 |
| T2D | Asian | Other | -1.0609978 | 0.28869092 | 0.57738184 |
| T2D | Asian | White | -1.5841392 | 0.11316204 | 0.43933621 |
| T2D | B/AA | Other | 0.39837874 | 0.69035103 | 0.82842123 |
| T2D | B/AA | White | -0.147501 | 0.88273658 | 0.88273658 |
| T2D | Other | White | -0.5372402 | 0.59110168 | 0.82842123 |

## References

Abraham Gnanadass, S., Divakar Prabhu, Y., & Valsala Gopalakrishnan, A. (2021). Association of metabolic and inflammatory markers with polycystic ovarian syndrome (PCOS): an update. Archives of Gynecology and Obstetrics, 303, 631–643.

Adashi, E. Y., Cibula, D., Peterson, M., & Azziz, R. (2023). The polycystic ovary syndrome: the first 150 years of study. F&S Reports, 4(1), 2–18.

Anagnostis, P., Tarlatzis, B. C., & Kauffman, R. P. (2018). Polycystic ovarian syndrome (PCOS): Long-term metabolic consequences. Metabolism, 86, 33–43.

Asuncion, M., Calvo, R. M., San Millán, J. L., Sancho, J., Avila, S., & Escobar-Morreale, H. c. F. (2000). A prospective study of the prevalence of the polycystic ovary syndrome in unselected Caucasian women from Spain. The Journal of Clinical Endocrinology & Metabolism, 85(7), 2434–2438.

Austin, P. C. (2011). An introduction to propensity score methods for reducing the effects of confounding in observational studies. Multivariate behavioral research, 46(3), 399–424.

Ávila, M. A. P. d., Bruno, R. V., Barbosa, F. C., Andrade, F. C. d., & Nardi, A. E. (2014). Polycystic ovary syndrome: implications of metabolic dysfunction. Revista do Colégio Brasileiro de Cirurgiões, 41, 106–110.

Azziz, R., Carmina, E., Chen, Z., Dunaif, A., Laven, J. S. E., Legro, R. S., Lizneva, D., Natterson-Horowtiz, B., Teede, H. J., & Yildiz, B. O. (2016). Polycystic ovary syndrome. Nature Reviews Disease Primers, 2(1), 16057. 10.1038/nrdp.2016.57

Balen, A. H., Morley, L. C., Misso, M., Franks, S., Legro, R. S., Wijeyaratne, C. N., Stener-Victorin, E., Fauser, B. C., Norman, R. J., & Teede, H. (2016). The management of anovulatory infertility in women with polycystic ovary syndrome: an analysis of the evidence to support the development of global WHO guidance. Human reproduction update, 22(6), 687–708.

Butt, A. S., & Devi, J. (2024). Polycystic ovary syndrome and nonalcoholic fatty liver disease. In Polycystic Ovary Syndrome (pp. 92–99). Elsevier.

Campbell, C. M., & Edwards, R. R. (2012). Ethnic differences in pain and pain management. Pain management, 2(3), 219–230.

Chaudhuri, A. (2023). Polycystic ovary syndrome: Causes, symptoms, pathophysiology, and remedies. Obesity Medicine, 39, 100480.

Cronin, L., Guyatt, G., Griffith, L., Wong, E., Azziz, R., Futterweit, W., Cook, D., & Dunaif, A. (1998). Development of a health-related quality-of-life questionnaire (PCOSQ) for women with polycystic ovary syndrome (PCOS). The Journal of Clinical Endocrinology & Metabolism, 83(6), 1976–1987.

Cui, N., Dai, T., Liu, Y., Wang, Y.-Y., Lin, J.-Y., Zheng, Q.-F., Zhu, D.-D., & Zhu, X.-W. (2024). Laryngopharyngeal reflux disease: Updated examination of mechanisms, pathophysiology, treatment, and association with gastroesophageal reflux disease. World journal of gastroenterology, 30(16), 2209.

Després, J.-P. (2012). Abdominal obesity and cardiovascular disease: is inflammation the missing link? Canadian Journal of Cardiology, 28(6), 642–652.

Drosdzol, A., Skrzypulec, V., Mazur, B., & Pawlińska-Chmara, R. (2007). Quality of life and marital sexual satisfaction in women with polycystic ovary syndrome. Folia histochemica et cytobiologica, 45(I), 93–97.

Elsenbruch, S., Hahn, S., Kowalsky, D., Öffner, A. H., Schedlowski, M., Mann, K., & Janssen, O. E. (2003). Quality of life, psychosocial well-being, and sexual satisfaction in women with polycystic ovary syndrome. The Journal of Clinical Endocrinology & Metabolism, 88(12), 5801–5807.

Escobar-Morreale, H. F., Luque-Ramírez, M., & González, F. (2011). Circulating inflammatory markers in polycystic ovary syndrome: a systematic review and metaanalysis. Fertility and sterility, 95(3), 1048–1058. e1042.

Eshre, T. R., & Group, A.-S. P. C. W. (2004). Revised 2003 consensus on diagnostic criteria and long-term health risks related to polycystic ovary syndrome. Fertility and sterility, 81(1), 19–25.

Georgescu, C. E. (2022). Polycystic ovary syndrome and nonalcoholic fatty liver disease. In Polycystic Ovary Syndrome (pp. 187–216). Elsevier.

Hadjiconstantinou, M., Mani, H., Patel, N., Levy, M., Davies, M., Khunti, K., & Stone, M. (2017). Understanding and supporting women with polycystic ovary syndrome: a qualitative study in an ethnically diverse UK sample. Endocrine connections, 6(5), 323–330.

Hahn, S., Janssen, O. E., Tan, S., Pleger, K., Mann, K., Schedlowski, M., Kimmig, R., Benson, S., Balamitsa, E., & Elsenbruch, S. (2005). Clinical and psychological correlates of quality-of-life in polycystic ovary syndrome. European journal of endocrinology, 153(6), 853–860.

Hampel, H., Abraham, N. S., & El-Serag, H. B. (2005). Meta-analysis: obesity and the risk for gastroesophageal reflux disease and its complications. Annals of internal medicine, 143(3), 199–211.

Jamieson, D. J., & Steege, J. F. (1996). The prevalence of dysmenorrhea, dyspareunia, pelvic pain, and irritable bowel syndrome in primary care practices. Obstetrics & Gynecology, 87(1), 55–58.

Kataria, S., & Ravindran, V. (2020). Electronic health records: a critical appraisal of strengths and limitations. Journal of the Royal College of Physicians of Edinburgh, 50(3), 262–268.

Kitzinger, C., & Willmott, J. (2002). ‘The thief of womanhood’: women’s experience of polycystic ovarian syndrome. Social science & medicine, 54(3), 349–361.

Kivimäki, M., Bartolomucci, A., & Kawachi, I. (2023). The multiple roles of life stress in metabolic disorders. Nature Reviews Endocrinology, 19(1), 10–27.

Kruse, C. S., Stein, A., Thomas, H., & Kaur, H. (2018). The use of electronic health records to support population health: a systematic review of the literature. Journal of medical systems, 42(11), 214.

Kumarendran, B., O’Reilly, M. W., Manolopoulos, K. N., Toulis, K. A., Gokhale, K. M., Sitch, A. J., Wijeyaratne, C. N., Coomarasamy, A., Arlt, W., & Nirantharakumar, K. (2018). Polycystic ovary syndrome, androgen excess, and the risk of nonalcoholic fatty liver disease in women: a longitudinal study based on a United Kingdom primary care database. PLoS medicine, 15(3), e1002542.

Lamping, D. L., Rowe, P., Clarke, A., Black, N., & Lessof, L. (1998). Development and validation of the menorrhagia outcomes questionnaire. BJOG: An International Journal of Obstetrics & Gynaecology, 105(7), 766–779.

Li, Y., Li, Y., Ng, E. H. Y., Stener-Victorin, E., Hou, L., Wu, T., Han, F., & Wu, X. (2011). Polycystic ovary syndrome is associated with negatively variable impacts on domains of health-related quality of life: evidence from a meta-analysis. Fertility and sterility, 96(2), 452–458.

Madden, J. M., Lakoma, M. D., Rusinak, D., Lu, C. Y., & Soumerai, S. B. (2016). Missing clinical and behavioral health data in a large electronic health record (EHR) system. Journal of the American Medical Informatics Association, 23(6), 1143–1149.

Maletzky, A., Böck, C., Tschoellitsch, T., Roland, T., Ludwig, H., Thumfart, S., Giretzlehner, M., Hochreiter, S., & Meier, J. (2022). Lifting hospital electronic health record data treasures: challenges and opportunities. JMIR Medical Informatics, 10(10), e38557.

McGowan, M. P. (2011). Polycystic ovary syndrome: a common endocrine disorder and risk factor for vascular disease. Current treatment options in cardiovascular medicine, 13(4), 289–301.

McHorney, C. A., Ware Johne, J., & Anastasiae, R. (1993). The MOS 36-Item Short-Form Health Survey (SF-36): II. Psychometric and clinical tests of validity in measuring physical and mental health constructs. Medical care, 31(3), 247–263.

Mousa, A., & Tay, C. T. (2023). Technical Report for the 2023 International Evidence-based Guideline for the Assessment and Management of Polycystic Ovary Syndrome: 2023 Update.

O’Brien, K., & Bosak, C. (2025). Primary Dysmenorrhea. In Medicinal Cannabis in Women’s Health: An Evidence-Based Clinician’s Guide (pp. 215–244). Springer.

Organization, W. H. (28 June 2023). Polycystic ovary syndrome. World Health Organization.

Osborn, O., & Olefsky, J. M. (2012). The cellular and signaling networks linking the immune system and metabolism in disease. Nature medicine, 18(3), 363–374.

Patel, S. (2018). Polycystic ovary syndrome (PCOS), an inflammatory, systemic, lifestyle endocrinopathy. The Journal of steroid biochemistry and molecular biology, 182, 27–36.

Penrod, N., Okeh, C., Velez Edwards, D. R., Barnhart, K., Senapati, S., & Verma, S. S. (2023). Leveraging electronic health record data for endometriosis research. Frontiers in Digital Health, 5, 1150687.

Portenoy, R. K., Ugarte, C., Fuller, I., & Haas, G. (2004). Population-based survey of pain in the United States: differences among white, African American, and Hispanic subjects. The journal of pain, 5(6), 317–328.

Qu, Z., Zhu, Y., Jiang, J., Shi, Y., & Chen, Z. (2013). The clinical characteristics and etiological study of nonalcoholic fatty liver disease in Chinese women with PCOS. Iranian journal of reproductive medicine, 11(9), 725.

Sai, B., & Mahaparale, S. (2024). Review on various disorders of the female reproductive system. Indian J. Applied & Pure Bio. *Vol,* 39(3), 1547–1556.

Sidra, S., Tariq, M. H., Farrukh, M. J., & Mohsin, M. (2019). Evaluation of clinical manifestations, health risks, and quality of life among women with polycystic ovary syndrome. PloS one, 14(10), e0223329.

Stein, I. F., & Leventhal, M. L. (1935). Amenorrhea associated with bilateral polycystic ovaries. American journal of obstetrics and gynecology., 29(2), 181–191. 10.1016/S0002-9378(15)30642-6

Teede, H., Deeks, A., & Moran, L. (2010). Polycystic ovary syndrome: a complex condition with psychological, reproductive and metabolic manifestations that impacts on health across the lifespan. BMC Medicine, 8(1), 41. 10.1186/1741-7015-8-41

Teede, H. J., Misso, M. L., Costello, M. F., Dokras, A., Laven, J., Moran, L., Piltonen, T., & Norman, R. J. (2018). Recommendations from the international evidence-based guideline for the assessment and management of polycystic ovary syndrome. Human reproduction, 33(9), 1602–1618.

Teede, H. J., Tay, C. T., Laven, J. J., Dokras, A., Moran, L. J., Piltonen, T. T., Costello, M. F., Boivin, J., Redman, L. M., & Boyle, J. A. (2023). Recommendations from the 2023 international evidence-based guideline for the assessment and management of polycystic ovary syndrome. European journal of endocrinology, 189(2), G43–G64.

Torres, D. M., & Harrison, S. A. (2016). Nonalcoholic Fatty Liver Disease: Clinical Features, Disease Modifiers, and Natural History. Alcoholic and Non-Alcoholic Fatty Liver Disease: Bench to Bedside, 183–194.

Tsilchorozidou, T., Overton, C., & Conway, G. S. (2004). The pathophysiology of polycystic ovary syndrome. Clinical endocrinology, 60(1), 1–17.

Walters, K. A., Gilchrist, R. B., Ledger, W. L., Teede, H. J., Handelsman, D. J., & Campbell, R. E. (2018). New perspectives on the pathogenesis of PCOS: neuroendocrine origins. Trends in Endocrinology & Metabolism, 29(12), 841–852.

Wu, P.-Y., Cheng, C.-W., Kaddi, C. D., Venugopalan, J., Hoffman, R., & Wang, M. D. (2016). –Omic and electronic health record big data analytics for precision medicine. IEEE Transactions on Biomedical Engineering, 64(2), 263–273.

Zawadri, J. (1992). Diagnostic criteria for polycystic ovary syndrome: towards a rational approach. Polycystic ovary syndrome. Current issues in endocrinology and metabolism.

Zhao, J., Papapetrou, P., Asker, L., & Boström, H. (2017). Learning from heterogeneous temporal data in electronic health records. Journal of biomedical informatics, 65, 105–119.

Zore, T., Joshi, N. V., Lizneva, D., & Azziz, R. (2017). Polycystic ovarian syndrome: long-term health consequences. Seminars in Reproductive Medicine,

